# Integrating HiTOP and RDoC Frameworks Part II: Shared and Distinct Biological Mechanisms of Externalizing and Internalizing Psychopathology

**DOI:** 10.1101/2025.02.05.25321732

**Authors:** Christal N. Davis, Yousef Khan, Sylvanus Toikumo, Zeal Jinwala, Dorret I. Boomsma, Daniel F. Levey, Joel Gelernter, Rachel L. Kember, Henry R. Kranzler

## Abstract

**Background:** The Hierarchical Taxonomy of Psychopathology (HiTOP) and Research Domain Criteria (RDoC) frameworks emphasize transdiagnostic and mechanistic aspects of psychopathology, respectively. We used a multi-omics approach to examine how externalizing (EXT), internalizing (INT), and shared EXT+INT liability map onto these models.

**Methods:** We conducted analyses across five RDoC units of analysis: genes, molecules, cells, circuits, and physiology. Using genome-wide association studies from the companion Part I article, we identified genes and tissue-specific expression patterns. We used drug repurposing analyses that integrate gene annotations to identify potential therapeutic targets and single-cell RNA sequencing data to implicate brain cell types. We then used magnetic resonance imaging data to examine brain regions and circuits associated with each psychopathology spectrum. Finally, we tested causal relationships between each spectrum and physical health conditions.

**Results:** Using five gene identification methods, EXT was associated with 1,759 genes, INT with 454 genes, and EXT+INT with 1,138 genes. Drug repurposing analyses identified potential therapeutic targets, including those that affect dopamine and serotonin pathways. Expression of EXT genes was enriched in GABAergic, cortical, and hippocampal neurons, while INT genes were more narrowly linked to GABAergic neurons. EXT+INT liability was associated with reduced gray matter volume in the amygdala and subcallosal cortex. Using Mendelian randomization, INT showed stronger causal effects on physical health—including chronic pain and cardiovascular diseases—than EXT.

**Conclusions:** Our findings revealed shared and distinct pathways underlying psychopathology. Integrating genomic insights with the RDoC and HiTOP frameworks advanced our understanding of mechanisms that underlie EXT and INT psychopathology.

## Introduction

The history of psychiatric nosology reflects an ongoing tension between the search for discrete categories of mental illness and the reality of complex, overlapping symptomatology. Although categorical classification systems like the Diagnostic and Statistical Manual of Mental Disorders (e.g., DSM-5) (American Psychiatric Association, 2013) provide a common language for diagnosis, they have faced criticism for their unreliability (Gordon & Heimberg, 2011) and the high degree of heterogeneity and comorbidity they yield (Borgogna, Owen, & Aita, 2024). At an aggregate level, there are more than ten million unique symptom combinations that can result in a diagnosed mental illness and almost two million ways to present with symptoms without meeting criteria for *any* diagnosis (Borgogna et al., 2024). This imprecision and variability hinders progress in psychiatric research, including efforts to develop targeted interventions.

Compounding these issues is the problem of multiple realizability, where a given psychiatric disorder can emerge from many biological, psychological, and environmental pathways (Kendler, 2022b). Thus, two individuals with depression may present with the same symptoms but differ in the mechanisms that underlie the disorder. This complexity makes it difficult to map psychiatric disorders onto biological etiologies, a challenge amplified by the “curse of polygenicity” (Kendler, 2022a, 2022b). Psychiatric disorders, among the most complex human traits, are influenced by thousands of genetic variants, each exerting only a small effect (Holland et al., 2020). The highly polygenic nature of these conditions increases the likelihood that many different combinations of genetic and environmental factors produce similar clinical presentations. Findings from recent genome-wide association studies (GWAS) highlight this complexity. Although GWAS have identified hundreds of loci associated with psychiatric disorders (Als et al., 2023; Trubetskoy et al., 2022), translating these findings into meaningful biological insights has been elusive.

Recent shifts toward dimensional and transdiagnostic frameworks, such as the Hierarchical Taxonomy of Psychopathology (HiTOP) and the Research Domain Criteria (RDoC), aim to address these limitations. HiTOP organizes psychopathology along hierarchical dimensions, from narrow symptoms to broad spectra, culminating in an overarching general psychopathology factor (*p*-factor) (Kotov et al., 2017). By contrast, RDoC focuses on identifying transdiagnostic mechanisms acting across multiple units of analysis, including genes, cells, circuits, and behaviors (Insel et al., 2010). These frameworks offer complementary approaches to conceptualizing and studying mental health, with both emphasizing shared and specific liabilities rather than rigid diagnostic categories.

The connection between mental and physical health further underscores the complexity and clinical relevance of these frameworks. Psychiatric disorders frequently co-occur with physical health conditions like chronic pain, cardiovascular disease, and other chronic illnesses, reflecting shared genetic etiologies (Davis et al., 2024; Lawrence et al., 2024). Furthermore, these conditions are clinically significant outcomes that conceptually align with externalizing (EXT) and internalizing (INT) psychopathology. For example, individuals who score higher on measures of negative valence report greater pain severity (Sambuco et al., 2022), while cardiovascular disease and other chronic illnesses are influenced by stress-related pathways and behavioral risk factors (e.g., smoking) tied to EXT and INT liability (Burke, Genuardi, Shappell, D’Agostino, & Magnani, 2017; Tully, Harrison, Cheung, & Cosh, 2016). Recently, it has been proposed that sensory processing be added to RDoC (Harrison, Kats, Williams, & Aziz-Zadeh, 2019), which has particular relevance for understanding pain as both a physical and psychological phenomenon. The considerable overlap between physical and mental health raises questions about how physical health conditions fit within psychopathology models like HiTOP, which tentatively incorporates a somatoform spectrum, and whether mechanisms identified in RDoC extend across both health domains.

Despite the potential that an integration of HiTOP and RDoC has to advance psychiatric nosology, significant gaps in that effort remain. Although HiTOP’s emphasis on shared psychopathology spectra can provide insight into the co-occurrence of EXT and INT problems, it offers no guidance on the mechanistic underpinnings of these spectra. RDoC complements HiTOP by emphasizing mechanisms, but its framework lacks the hierarchical structure needed to reconcile overlapping yet distinct components of psychopathology. Bridging these complementary frameworks could accelerate etiological research and facilitate the development of novel treatments for psychiatric disorders (Michelini, Palumbo, DeYoung, Latzman, & Kotov, 2021).

Here, we integrate the complementary perspectives of HiTOP and RDoC by examining genetic associations both hierarchically, as suggested by HiTOP, and across multiple units of analysis, as emphasized by RDoC. Specifically, we examine mechanisms across genes, molecules, cells, circuits, physiology, and behavior to determine whether associations are unique to each spectrum or reflect common mechanisms of both. Building on insights into the genetic architecture of these spectra provided in the Part I companion article, this study offers a novel integration aimed at deepening our understanding of the genetic architecture of EXT and INT psychopathology and informing more precise approaches to diagnosis, classification, and treatment.

## Methods

### Genome-Wide Association Studies

This study uses summary statistics from GWAS conducted in the companion Part I article for EXT, INT, and their shared liability (EXT+INT). The GWAS were derived from genomic structural equation models (gSEM) of 16 clinical and subclinical EXT (e.g., substance use disorders and risky behaviors) and INT (e.g., mood, anxiety, and wellbeing) traits. The GWAS results serve as the foundation for all downstream analyses in this paper, and details on the GWAS methods employed can be found in Part I.

### Unit of Analysis: Genes

#### Gene Expression and Enrichment

We identified genes associated with EXT, INT, and their shared liability (EXT+INT) using multiple approaches. First, we performed gene-based tests using MAGMA (de Leeuw, Mooij, Heskes, & Posthuma, 2015), which aggregates association signals from genetic variants to genes based on their position. Second, we mapped additional genes based on functional effects, including expression quantitative trait loci (eQTLs), which link genetic variation to gene activity, and chromatin interaction data, which identify physical connections between genomic regions. We then examined the expression of associated genes across developmental stages and within specific brain regions using data from BrainSpan (Li et al., 2018) and GTEx v8 (The GTEx Consortium et al., 2020).

#### Transcriptome-Wide Association Studies

To prioritize potential causal genes, we conducted transcriptome-wide association studies (TWAS) using two complementary methods. TWAS identifies genes whose predicted expression levels are associated with a trait by integrating GWAS data with gene expression profiles to help pinpoint genes that may play a causal role in the biological pathways underlying the GWAS trait. First, we used S-MultiXcan (Barbeira et al., 2019) to simultaneously integrate gene expression data across 13 brain tissues. Next, we used S-PrediXcan (Barbeira et al., 2018) to focus specifically on data from the frontal and temporal cortices of psychiatric cases and controls (Gandal et al., 2018; Jourdon, Scuderi, Capauto, Abyzov, & Vaccarino, 2021). We applied a Bonferroni correction to account for multiple testing.

### Unit of Analysis: Molecules

To identify druggable targets—genes that encode proteins that can be modulated by existing drugs or are predicted to be viable candidates for therapeutic development—we mapped genes associated with EXT, INT, and EXT+INT to databases of known gene-drug interactions using the Drug-Gene-Interaction Database (Freshour et al., 2021). We included genes that interact with currently approved medications and investigational compounds, prioritizing targets supported by multiple lines of genetic evidence, such as chromatin interactions (which identify physical connections between genetic regions) and eQTL analyses (which link genetic variation to gene activity). For EXT and INT, we focused on genes associated with one spectrum but not the other, aiming to identify drugs that could provide targeted therapeutic options based on biological mechanisms specific to EXT or INT.

### Unit of Analysis: Cells

We examined genetic effects on brain cell types using single-cell RNA sequencing (scRNA-seq) datasets from 15 human brain cell expression profiles (Darmanis et al., 2015; Habib et al., 2017; La Manno et al., 2016; Watanabe, Umićević Mirkov, de Leeuw, van den Heuvel, & Posthuma, 2019). Independently associated cell types were identified using three-step conditional analyses. First, we identified significant cell types within each dataset. Next, we performed within-dataset conditional analyses to identify independent associations among correlated cell types. Finally, we conducted cross-dataset conditional analyses to assess whether associations reflected shared genetic signals across the datasets.

### Unit of Analysis: Circuits

To examine how EXT, INT, and EXT+INT relate to brain structures and connectivity, we used BrainXcan (Liang et al., 2022), which links genetic data to 327 imaging-derived phenotypes (IDPs) from structural and diffusion magnetic resonance imaging (MRI) scans. Effect sizes and p-values were adjusted using linkage disequilibrium (LD) block-based permutation to account for inflation in test statistics that can arise from the correlations among nearby genetic variants. We also applied a Bonferroni correction to account for multiple testing.

### Unit of Analysis: Physiology

To evaluate potentially causal impacts on 15 physical health traits, we conducted Generalized Summary-data-based Mendelian Randomization (GSMR) analyses (Zhu et al., 2018). GSMR estimates the potential causal effects of exposures (i.e., EXT, INT, or EXT+INT) on outcomes (i.e., physical health traits) by leveraging GWAS summary statistics and treating genetic variants as instrumental variables. We based instrument selection on a genome-wide significance threshold of *p* < 5×10^-8^ to ensure robust genetic associations. To address horizontal pleiotropy, which violates a major assumption of Mendelian randomization analyses requiring that instruments influence the outcome solely through the exposure, we applied the heterogeneity in dependent instruments (HEIDI)-outlier method (Zhu et al., 2018). This excludes genetic variants with evidence of such pleiotropic effects to reduce bias in causal estimates. We applied a Bonferroni correction to account for multiple testing.

Physical health traits were chosen from four domains with strong empirical and theoretical links to psychopathology: (1) pain, (2) general health, (3) cardiovascular disease, and (4) other chronic illnesses (Isvoranu et al., 2021; Lawrence et al., 2024; Waszczuk et al., 2023; Zhang et al., 2021). The pain domain comprised GWAS of pain intensity (Toikumo et al., 2024), multisite chronic pain (Johnston et al., 2019), and back pain (Freidin et al., 2019). General health indices included summary statistics from GWAS of longstanding illness, disability, or infirmity; hospitalization; and age at death conducted in the UK Biobank (http://www.nealelab.is/uk-biobank/). Cardiovascular disease GWAS comprised five traits: (1) heart failure (Zhou et al., 2022), (2) stroke (Zhou et al., 2022), (3) myocardial infarction (Hartiala et al., 2021), (4) hypertension (http://www.nealelab.is/uk-biobank/), and (5) abdominal aortic aneurysm (Zhou et al., 2022). Finally, we selected four GWAS of other chronic illnesses: (1) type 2 diabetes (Mahajan et al., 2022), (2) inflammatory bowel disease (IBD) (Liu et al., 2023), (3) chronic obstructive pulmonary disease (Zhou et al., 2022), and (4) asthma (Zhou et al., 2022).

## Results

### Unit of Analysis: Genes

#### Gene Expression and Enrichment

Using MAGMA’s genome-wide gene-based test, we identified 326 genes associated with EXT but not INT (Supplementary Table 1; e.g., *CADM2* and *FTO*). Although not differentially expressed during any developmental period, EXT-specific genes were differentially expressed in four brain tissues: the hippocampus, amygdala, putamen, and caudate (Supplementary Figure 1). A gene set related to mRNA binding was the only significant association (Supplementary Table 2). There were 31 genes associated with INT but not EXT (Supplementary Table 3; e.g., *CCDC68*). The INT-specific genes were not differentially expressed in any developmental stages or tissue types, and no gene sets were significant. Of the genes associated with EXT+INT (Supplementary Table 4), 29 were also identified by both first-order factors (e.g., *DRD2*, *NCAM1*, and *DCC*). Gene expression for EXT+INT was significantly upregulated during early and early mid-prenatal periods and downregulated during early childhood. EXT+INT genes were upregulated in 10 brain regions, including the frontal cortex, amygdala, anterior cingulate cortex, and hippocampus (Supplementary Figure 2), but no gene sets were significant.

#### Transcriptome-Wide Association Studies

Using S-MultiXcan to predict effects on gene expression across 13 brain tissues revealed 352 significant genes for EXT, 141 for INT, and 238 for EXT+INT (Figure 1, Supplementary Tables 5-7, and Supplementary Figure 3). TWAS using PsychENCODE data identified 207 genes for EXT, 52 for INT, and 124 for EXT+INT (Supplementary Tables 8-10 and Supplementary Figure 4). Forty-five genes were identified in both TWAS for EXT, 21 for INT, and 36 for EXT+INT (Supplementary Figures 5-7).

**Figure 1.**
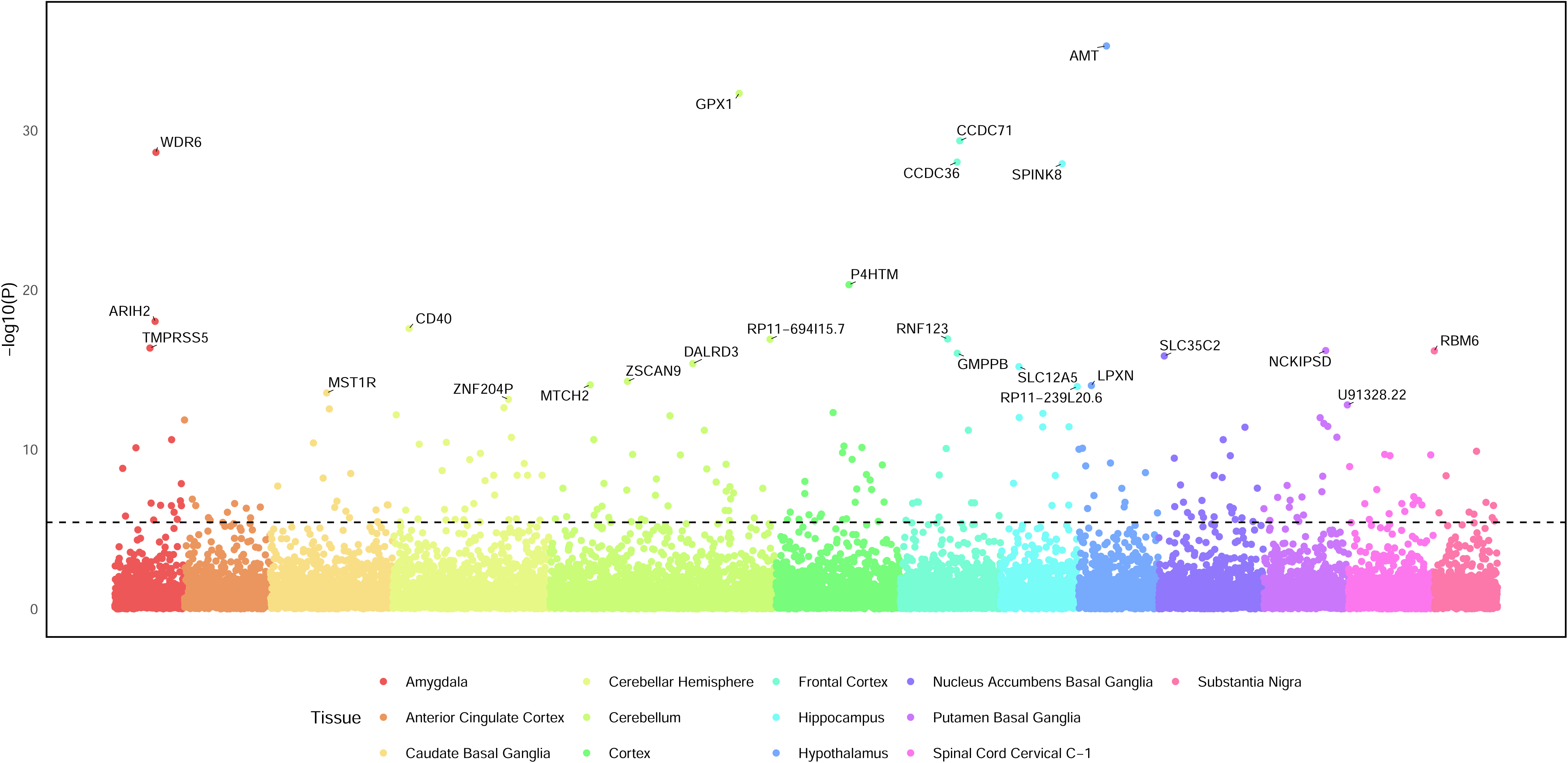
Transcriptome wide association study (TWAS) for the externalizing and internalizing (EXT+INT) factor across 13 brain tissues. Gene names for the top 25 significant associations are annotated. Significance was determined using a Bonferroni-adjusted p-value of 3.73 × 10^-6^ (0.05/13,406 tests). The dashed line at 5.43 indicates the significance level (-log10(3.73 x 10^-6^). A total of 236 associations were significant after multiple testing correction. Full TWAS results for all the factors can be found in Supplementary Tables 5-10.

#### Integration Across Gene Identification Methods

Collectively, across all five gene identification methods (MAGMA, chromatin interactions, eQTLs, S-MultiXcan, and S-PrediXcan), we identified 1,759 genes associated with EXT, 454 with INT, and 1,138 with EXT+INT (Supplementary Figure 8). For EXT, 20.41% of associated genes were identified using more than one method, including 7 genes—*AS3MT*, *BTN3A2*, *KHK*, *NOB1*, *RPGRIP1L*, *SMIM19*, *ZKSCAN3*—identified by all five methods. For INT, 20.04% of genes were identified by more than one method, and *QRICH1* was identified by all five methods. For EXT+INT, 23.20% of genes were identified by more than one method, and 3 genes—*ZKSCAN8*, *TMA7*, and *TREX1*—were identified by all five methods (Figure 2).

**Figure 2.**
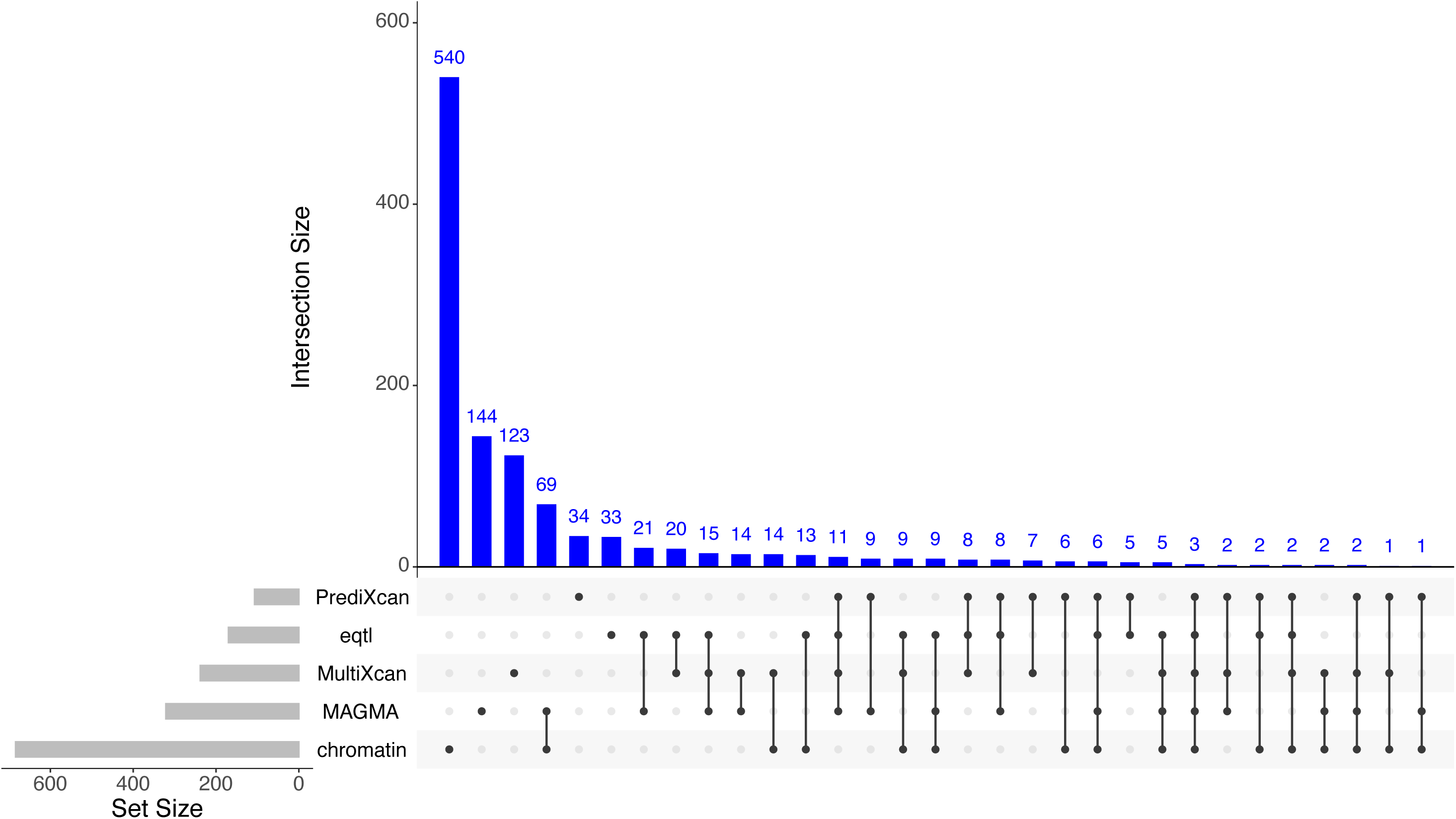
UpSet plot of genes identified for the shared liability to externalizing and internalizing (EXT+INT).

### Unit of Analysis: Molecules

Among the genes identified for EXT, 52 druggable targets specific to EXT were identified by at least two methods. The 52 genes yielded 492 drug-gene interactions (Supplementary Table 11), including with dextroamphetamine, phenobarbital, baclofen, naltrexone, naloxone, and methadone. Interactions with medications from anti-migraine, anti-inflammatory, and anticonvulsant therapeutic classes (e.g., topiramate and lamotrigine) were also identified. Most identified drugs (64.84%) have regulatory approval from the U.S. Food and Drug Administration (FDA).

Of the genes associated with INT, 7 druggable targets specific to INT were identified by at least two methods, yielding 292 drug-gene interactions (Supplementary Table 12). Drug targets included antidepressants and antipsychotics. Unlike for EXT, most drugs identified for INT (82.33%) are not currently approved by the FDA. For EXT+INT, 47 of the identified genes were druggable targets implicated by more than one method. Of the 460 drug-gene interactions identified (Supplementary Table 13), many were also present in the INT or EXT results. Most of these drugs (75.52%) are not currently approved by the FDA.

### Unit of Analysis: Cells

EXT was significantly associated with dopaminergic and GABAergic neurons and neuroblasts from embryonic brain samples, human cortical neurons and hybrid cells, and pyramidal neurons from the cornu ammonis (CA1) region of the hippocampus. After conditional analyses, there were independent significant associations for GABAergic, cortical, and hippocampal neurons (Supplementary Figure 9). The only significant cell-type association for INT was with GABAergic neurons, though it was not independently significant after conditional analyses. EXT+INT showed significant associations with dopaminergic, GABAergic, and cortical neurons, though these associations were also not independently significant.

### Unit of Analysis: Circuits

Twenty brain IDPs were associated with EXT (Supplementary Figures 10-11 and Supplementary Table 14), including greater gray matter volumes in the thalamus, caudate nuclei, and occipital pole, and lower volumes in the ventral striatum and amygdala. There were also significant associations with intra-cellular volume fraction or orientation dispersion indices (ODI) in the medial lemniscus, cerebral peduncle, and middle cerebellar peduncle. INT was significantly associated with lower gray matter volume in the subcallosal cortex (Supplementary Figures 12-13 and Supplementary Table 15). EXT+INT showed negative associations with gray matter volume in the amygdala and subcallosal cortex (Figure 3), positive associations with ODI in the medial lemniscus and cerebellar peduncle, and negative associations with ODI in the external capsule (Supplementary Table 16 and Supplementary Figures 14 and 15).

**Figure 3.**
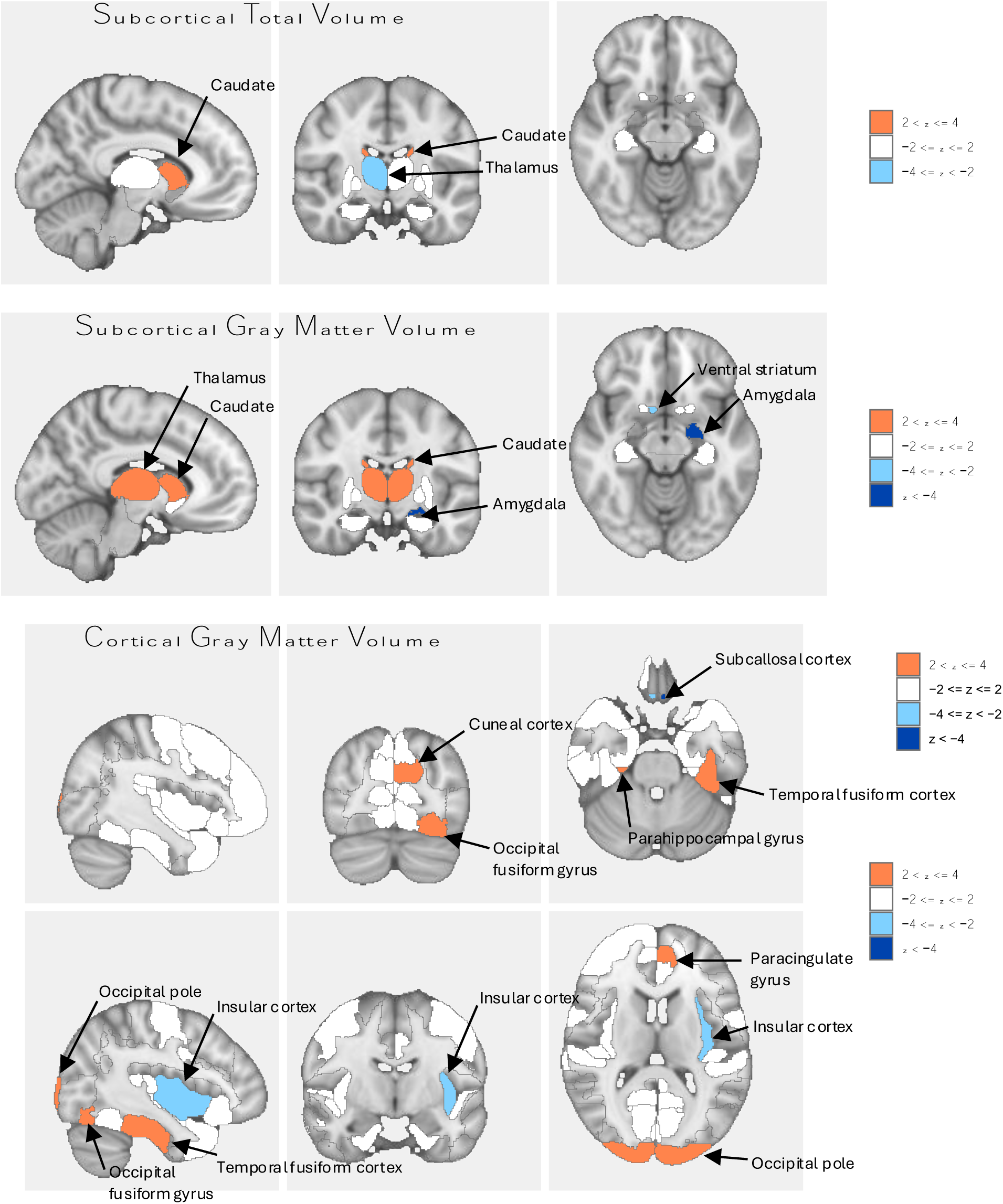
Brain region associations for the externalizing and internalizing (EXT+INT) factor. Associations shown are for image-derived phenotypes from structural magnetic resonance imaging. Full results are in Supplementary Figures 10-15 and Supplementary Tables 14-16. Blue colors represent reduced volume, and orange represents increased volume.

### Unit of Analysis: Physiology

EXT had significant positive causal effects on all physical health traits, except age at death and IBD. INT was causally associated with all traits except age at death and abdominal aortic aneurysm. INT had protective effects on IBD (*b_xy_* = -0.32, SE = 0.09, *p* = 0.0004), but stronger positive associations than EXT with all pain phenotypes, all cardiovascular diseases, and three of four other chronic illnesses (Figure 4). Like EXT, EXT+INT had positive causal effects on all physical health traits except age at death and IBD (Supplementary Table 17).

**Figure 4.**
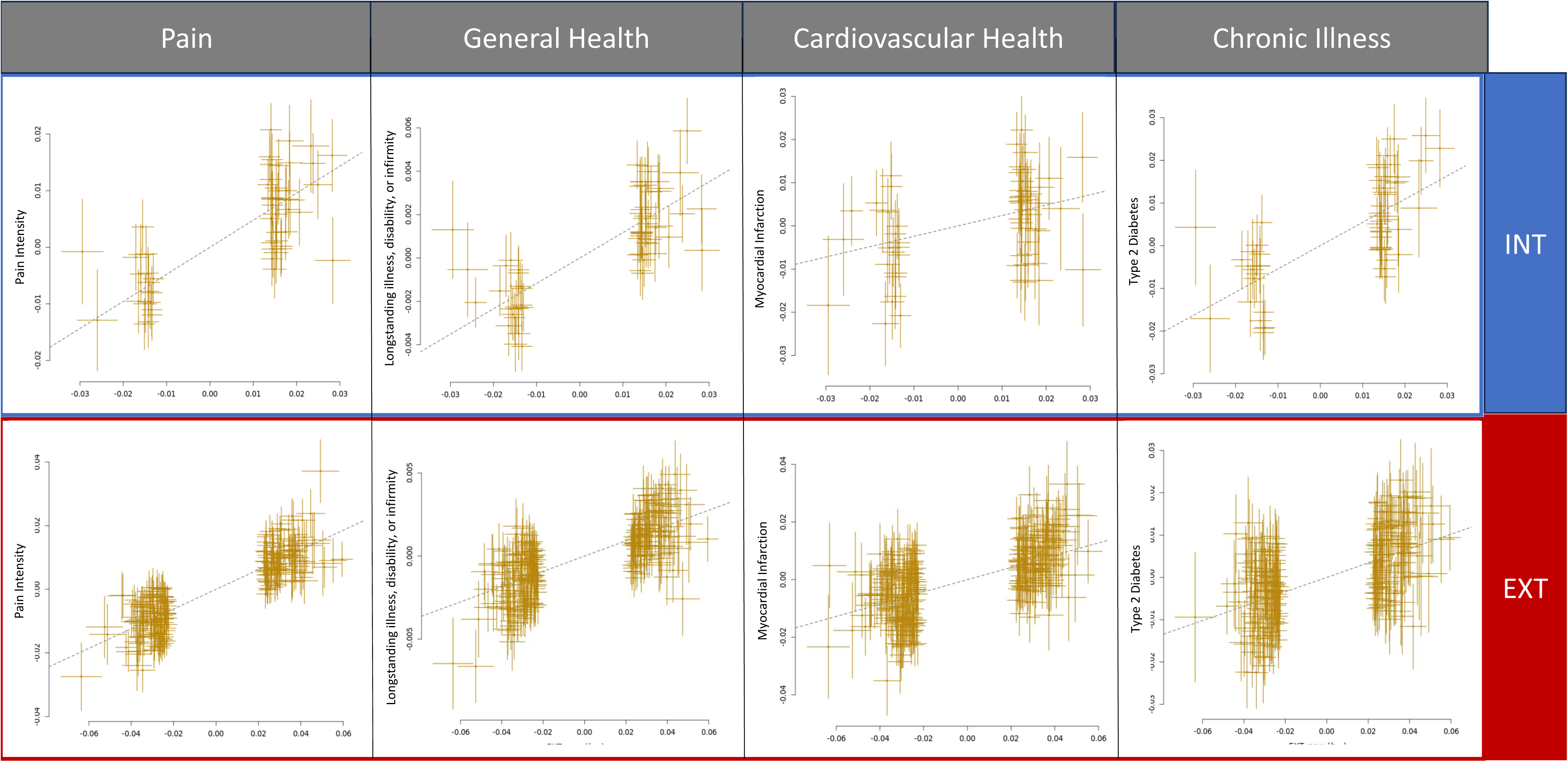
Representative results of generalized summary-data-based Mendelian Randomization (GSMR) across four domains of physical health. Pain intensity results: INT: *b_xy_* = 0.48, SE = 0.04, *p* = 1.11E-28; EXT: *b_xy_* = 0.31, SE = 0.01, *p* = 1.44E-155. Longstanding illness, disability, or infirmity results: INT: OR = 1.12, SE = 0.01, *p* = 9.83E-33; EXT: OR = 1.05, SE = 0.002, *p* = 2.27E-75. Myocardial infarction results: INT: OR = 1.27, SE = 0.07, *p* = 0.0003; EXT: OR = 1.24, SE = 0.02, *p* = 6.32E-38. Type 2 Diabetes results: INT: OR = 1.72, SE = 0.06, *p* = 7.28E-20; EXT: OR = 1.19, SE = 0.02, *p* = 6.94E-28. All results depicted are significant at a Bonferroni corrected p-value of 0.001. In all analyses, INT/EXT is the exposure, and the physical health trait is the outcome. INT = internalizing, EXT = externalizing, OR = odds ratio. Full results can be found in Supplementary Table 17.

## Discussion

Our findings demonstrate the value of integrating the HiTOP and RDoC frameworks to advance our understanding of the biological underpinnings of EXT and INT psychopathology. HiTOP’s hierarchical structure contextualizes the co-occurrence of psychopathology across levels and the unique features at each level. In turn, RDoC complements HiTOP with a focus on transdiagnostic mechanisms. By combining the two frameworks, our results provide insights that each alone cannot fully capture. For example, although HiTOP conceptualizes overarching patterns of comorbidity, it does not directly address mechanisms. Conversely, RDoC’s mechanistic approach lacks a framework to reconcile the shared and distinct mechanisms among overlapping psychopathology spectra. Genetics provides a promising avenue for bridging these models, offering a lens to identify shared and unique biological mechanisms across EXT and INT and to connect dimensions of psychopathology to their underlying etiology.

In line with HiTOP’s hierarchical model, which posits that higher-order psychopathology factors emerge from shared features between lower-level spectra, we identified etiologic factors that contribute to the overlap between EXT and INT psychopathology. Genes like *DRD2, NCAM1,* and *DCC* mapped to both spectra. *DRD2* encodes the dopamine D2 receptor and is implicated in traits related to reward processing and impulse control, including substance use and mood disorders (Friligkou et al., 2024; Levey et al., 2023; Meng et al., 2024). *NCAM1* is involved in neural cell adhesion and synaptic plasticity, processes critical for cognitive and emotional regulation (Conboy, Bisaz, Markram, & Sandi, 2010), while *DCC* encodes a receptor involved in nervous system development and has been linked to psychiatric disorders (Friligkou et al., 2024) and cognitive performance (Williams, Labouret, Wolfram, Peyre, & Ramus, 2023). These genes have been shown to have pleiotropic effects across multiple forms of psychopathology (Lam et al., 2022).

The EXT+INT liability also implicated neural structural differences, such as lower gray matter volume in the amygdala and subcallosal cortex. Lower left amygdala volume has been shown to mediate the relation between childhood threat exposure and the development of EXT and INT symptoms (Picci et al., 2022). Similarly, lower left subcallosal cortical volume is a potential mediator of associations between personality, emotional states, and psychiatric disorders (Wendt et al., 2021). EXT+INT liability was also associated with white matter fiber differences, potentially reflecting disruptions in neural communication pathways that contribute to psychopathology (Kraguljac, Guerreri, Strickland, & Zhang, 2023).

Alongside shared etiologic factors, we identified genes with specific associations to each spectrum: *CADM2* and *FTO* were uniquely associated with EXT traits, suggesting the involvement of pathways related to impulsivity, risk-taking, and reward sensitivity—key components of RDoC’s cognitive and positive valence systems. *CADM2* encodes a cell adhesion molecule and has been associated with impulsivity and risky behaviors (e.g., substance use; Sanchez-Roige et al., 2023), while the *FTO* gene has been implicated in obesity (Huang, Chen, & Wang, 2023) and substance use (Hatoum et al., 2023; Kember et al., 2023). For INT, *CCDC68*, involved in cellular structure (Huang et al., 2017), exhibited specificity. This gene has been associated with several INT traits, including depression (Meng et al., 2024) and neuroticism (Baselmans et al., 2019).

Building on the gene associations, drug repurposing analyses identified potential druggable targets specific to each spectrum, including several novel compounds for INT, highlighting opportunities for precision treatment. Genes associated with each spectrum also showed specific expression patterns in brain cell types. EXT genes showed enriched expression in GABAergic, cortical, and pyramidal hippocampal neurons, cell types that align with RDoC constructs related to cognitive control and acute threat. In contrast, INT genes were more narrowly associated with GABAergic neurons involved in inhibitory signaling, consistent with RDoC’s negative valence systems, particularly the acute threat construct.

Genetic liability for both the EXT and INT spectra had downstream effects on physical health outcomes. INT liability was causally associated with chronic pain, cardiovascular disease, and other chronic illnesses, consistent with previous research supporting causal links between INT, localized pain (Williams et al., 2022; Yao et al., 2023), and disease outcomes (Mulugeta, Zhou, King, & Hyppönen, 2020). These findings suggest that the negative valence systems implicated in INT psychopathology may extend to somatic outcomes, highlighting shared biological pathways between psychiatric and physical health (Lawrence et al., 2024). EXT liability also exhibited associations, albeit generally weaker, with adverse health outcomes, indicating that liability for both spectra contributes to physical health risk.

The combination of shared and distinct mechanisms reinforces the need for an integrated HiTOP-RDoC framework where shared liabilities are contextualized alongside spectrum-specific biological underpinnings. Traditional diagnostic systems have been criticized for their categorical approach, which fails to account for substantial disorder-level heterogeneity and comorbidity. By integrating HiTOP and RDoC, we propose a dimensional-mechanistic framework that identifies shared pathways and distinguishes spectrum-specific mechanisms. This approach offers a cohesive framework for aligning transdiagnostic dimensions with biological mechanisms, paving the way for a more precise and biologically informed psychiatric nosology.

### Limitations

The use of summary statistics from large GWAS, comprehensive analyses across multiple RDoC units, and an innovative integration of HiTOP and RDoC frameworks using genomics are among the study’s strengths. However, findings should be interpreted in the context of several limitations. First, Mendelian randomization makes several key assumptions that influence the validity of causal inferences: (1) instrumental variables must be robustly associated with the exposure, (2) there must be no direct pathway from the instruments to the outcome other than through the exposure (i.e., no horizontal pleiotropy), and (3) instruments must not be associated with confounders of the exposure-outcome relationship. We implemented methods to address the first two assumptions. First, we selected instruments associated at *p* < 5×10^-8^ to ensure statistical rigor, addressing the first assumption. To address assumption two, we used the HEIDI-outlier method to detect and exclude genetic instruments with evidence of pleiotropic effects. However, because assumption three is particularly challenging to address, we cannot rule out the possibility of confounding, and the results should be interpreted cautiously. Additionally, the cross-sectional nature of the GWAS data limits our ability to make developmental inferences, as data are not available to identify temporal patterns of genetic and environmental mechanisms. Finally, our analyses were limited to individuals genetically similar to Europeans. Expanding future studies to additional populations is crucial both to ensure the replicability of these results and to extend their generalizability.

### Conclusions

Psychiatry has long grappled with the complexity of psychiatric symptomatology and the considerable challenge of linking clinical presentations to their underlying biological mechanisms. By integrating HiTOP’s dimensional approach with RDoC’s mechanistic focus, we demonstrate how research that bridges these frameworks can address limitations of our traditional diagnostic system. This integrative approach moves beyond rigid diagnostic categories to provide a model that can account for both shared and spectrum-specific pathways of psychopathology. A combined HiTOP-RDoC framework has the potential to address enduring challenges in psychiatry, such as the heterogeneity of psychiatric disorders and the numerous biological and environmental pathways that yield similar symptoms. This integration provided unified insights across genetic, neural, and clinical domains, which can ultimately refine psychiatric nosology, guide therapeutic development, and advance precision psychiatry.

## Data Availability

GWAS Summary Statistics used for the present study can be accessed at the following locations:
UK Biobank (http://www.nealelab.is/uk-biobank/), dbGaP accession phs001672 for Million Veteran Program (https://www.ncbi.nlm.nih.gov/projects/gap/cgi-bin/study.cgi?study_id=phs001672), iPSYCH (https://ipsych.dk/en/research/downloads/),
Psychiatric Genetics Consortium (https://pgc.unc.edu/for-researchers/download-results/), the Social Science Genetic Association Consortium (https://thessgac.com/), GWAS catalog
(https://www.ebi.ac.uk/gwas/publications/28979981; https://www.ebi.ac.uk/gwas/publications/33532862), Gelernter Lab website
(https://medicine.yale.edu/lab/gelernter/stats/), Global Biobank Meta-analysis Initiative
(https://www.globalbiobankmeta.org/resources), and the Diabetes Genetics Replication and
Meta-Analysis Consortium (https://diagram-consortium.org/downloads.html). Once accepted
for publication, summary statistics for the externalizing, internalizing, and second-order
psychopathology factors will be made publicly available.

## References

Als, T. D., Kurki, M. I., Grove, J., Voloudakis, G., Therrien, K., Tasanko, E., … Børglum, A. D. (2023). Depression pathophysiology, risk prediction of recurrence and comorbid psychiatric disorders using genome-wide analyses. Nature Medicine, 29(7), 1832–1844. doi:10.1038/s41591-023-02352-1

American Psychiatric Association. (2013). *Diagnostic and statistical manual of mental disorders: DSM-5* (Fifth edition. ed.). Washington, D.C.: American Psychiatric Publishing.

Barbeira, A. N., Dickinson, S. P., Bonazzola, R., Zheng, J., Wheeler, H. E., Torres, J. M., … Visualization—Ucsc Genomics Institute, U. o. C. S. C. (2018). Exploring the phenotypic consequences of tissue specific gene expression variation inferred from GWAS summary statistics. Nature Communications, 9(1), 1825. doi:10.1038/s41467-018-03621-1

Barbeira, A. N., Pividori, M., Zheng, J., Wheeler, H. E., Nicolae, D. L., & Im, H. K. (2019). Integrating predicted transcriptome from multiple tissues improves association detection. PLOS Genetics, 15(1), e1007889. doi:10.1371/journal.pgen.1007889

Baselmans, B. M. L., Jansen, R., Ip, H. F., van Dongen, J., Abdellaoui, A., van de Weijer, M. P., … Bartels, M. (2019). Multivariate genome-wide analyses of the well-being spectrum. Nat Genet, 51(3), 445–451. doi:10.1038/s41588-018-0320-8

Borgogna, N. C., Owen, T., & Aita, S. L. (2024). The absurdity of the latent disease model in mental health: 10,130,814 ways to have a DSM-5-TR psychological disorder. Journal of Mental Health, 33(4), 451–459. doi:10.1080/09638237.2023.2278107

Burke, G. M., Genuardi, M., Shappell, H., D’Agostino, R. B., & Magnani, J. W. (2017). Temporal associations between smoking and cardiovascular disease, 1971 to 2006 (from the Framingham Heart Study). The American Journal of Cardiology, 120(10), 1787–1791. doi:10.1016/j.amjcard.2017.07.087

Conboy, L., Bisaz, R., Markram, K., & Sandi, C. (2010). Role of NCAM in Emotion and Learning. In V. Berezin (Ed.), Structure and Function of the Neural Cell Adhesion Molecule NCAM (pp. 271–296). New York, NY: Springer New York.

Darmanis, S., Sloan, S. A., Zhang, Y., Enge, M., Caneda, C., Shuer, L. M., … Quake, S. R. (2015). A survey of human brain transcriptome diversity at the single cell level. Proceedings of the National Academy of Science USA, 112(23), 7285–7290. doi:10.1073/pnas.1507125112

Davis, C. N., Toikumo, S., Hatoum, A. S., Khan, Y., Pham, B. K., Pakala, S. R., … Kranzler, H. R. (2024). Multivariate, multi-omic analysis in 799,429 individuals identifies 134 loci associated with somatoform traits. medRxiv, 2024.2007.2029.24310991. doi:10.1101/2024.07.29.24310991

de Leeuw, C. A., Mooij, J. M., Heskes, T., & Posthuma, D. (2015). MAGMA: Generalized gene-set analysis of GWAS data. PLOS Computational Biology, 11(4), e1004219. doi:10.1371/journal.pcbi.1004219

Freidin, M. B., Tsepilov, Y. A., Palmer, M., Karssen, L. C., Group, C. M. W., Suri, P., … Williams, F. M. K. (2019). Insight into the genetic architecture of back pain and its risk factors from a study of 509,000 individuals. PAIN, 160(6), 1361–1373. doi:10.1097/j.pain.0000000000001514

Freshour, S. L., Kiwala, S., Cotto, K. C., Coffman, A. C., McMichael, J. F., Song, J. J., … Wagner, A. H. (2021). Integration of the Drug–Gene Interaction Database (DGIdb 4.0) with open crowdsource efforts. Nucleic Acids Research, 49(D1), D1144–D1151. doi:10.1093/nar/gkaa1084

Friligkou, E., Løkhammer, S., Cabrera-Mendoza, B., Shen, J., He, J., Deiana, G., … Polimanti, R. (2024). Gene discovery and biological insights into anxiety disorders from a large-scale multi-ancestry genome-wide association study. Nature Genetics, 56(10), 2036–2045. doi:10.1038/s41588-024-01908-2

Gandal, M. J., Zhang, P., Hadjimichael, E., Walker, R. L., Chen, C., Liu, S., … Abyzov, A. (2018). Transcriptome-wide isoform-level dysregulation in ASD, schizophrenia, and bipolar disorder. Science, 362(6420), eaat8127. doi:10.1126/science.aat8127

Gordon, D., & Heimberg, R. G. (2011). Reliability and validity of DSM-IV generalized anxiety disorder features. Journal of Anxiety Disorders, 25(6), 813–821. doi:10.1016/j.janxdis.2011.04.001

Habib, N., Avraham-Davidi, I., Basu, A., Burks, T., Shekhar, K., Hofree, M., … Regev, A. (2017). Massively parallel single-nucleus RNA-seq with DroNc-seq. Nature Methods, 14(10), 955–958. doi:10.1038/nmeth.4407

Harrison, L. A., Kats, A., Williams, M. E., & Aziz-Zadeh, L. (2019). The importance of sensory processing in mental health: A proposed addition to the Research Domain Criteria (RDoC) and suggestions for RDoC 2.0. Frontiers in Psychology, 10. doi:10.3389/fpsyg.2019.00103

Hartiala, J. A., Han, Y., Jia, Q., Hilser, J. R., Huang, P., Gukasyan, J., … Allayee, H. (2021). Genome-wide analysis identifies novel susceptibility loci for myocardial infarction. European Heart Journal, 42(9), 919–933. doi:10.1093/eurheartj/ehaa1040

Hatoum, A. S., Colbert, S. M. C., Johnson, E. C., Huggett, S. B., Deak, J. D., Pathak, G., … Agrawal, A. (2023). Multivariate genome-wide association meta-analysis of over 1 million subjects identifies loci underlying multiple substance use disorders. Nature Mental Health, 1(3), 210–223. doi:10.1038/s44220-023-00034-y

Holland, D., Frei, O., Desikan, R., Fan, C.-C., Shadrin, A. A., Smeland, O. B., … Dale, A. M. (2020). Beyond SNP heritability: Polygenicity and discoverability of phenotypes estimated with a univariate Gaussian mixture model. PLOS Genetics, 16(5), e1008612. doi:10.1371/journal.pgen.1008612

Huang, C., Chen, W., & Wang, X. (2023). Studies on the fat mass and obesity-associated (FTO) gene and its impact on obesity-associated diseases. Genes & Diseases, 10(6), 2351–2365. doi:10.1016/j.gendis.2022.04.014

Huang, N., Xia, Y., Zhang, D., Wang, S., Bao, Y., He, R., … Chen, J. (2017). Hierarchical assembly of centriole subdistal appendages via centrosome binding proteins CCDC120 and CCDC68. Nature Communications, 8(1), 15057. doi:10.1038/ncomms15057

Insel, T., Cuthbert, B., Garvey, M., Heinssen, R., Pine, D. S., Quinn, K., … Wang, P. (2010). Research Domain Criteria (RDoC): Toward a new classification framework for research on mental disorders. American Journal of Psychiatry, 167(7), 748–751. doi:10.1176/appi.ajp.2010.09091379

Isvoranu, A. M., Abdin, E., Chong, S. A., Vaingankar, J., Borsboom, D., & Subramaniam, M. (2021). Extended network analysis: From psychopathology to chronic illness. BMC Psychiatry, 21(1), 119. doi:10.1186/s12888-021-03128-y

Johnston, K. J. A., Adams, M. J., Nicholl, B. I., Ward, J., Strawbridge, R. J., Ferguson, A., … Smith, D. J. (2019). Genome-wide association study of multisite chronic pain in UK Biobank. PLOS Genetics, 15(6), e1008164. doi:10.1371/journal.pgen.1008164

Jourdon, A., Scuderi, S., Capauto, D., Abyzov, A., & Vaccarino, F. M. (2021). PsychENCODE and beyond: Transcriptomics and epigenomics of brain development and organoids. Neuropsychopharmacology, 46(1), 70–85. doi:10.1038/s41386-020-0763-3

Kember, R. L., Vickers-Smith, R., Zhou, H., Xu, H., Jennings, M., Dao, C., … Kranzler, H. R. (2023). Genetic underpinnings of the transition from alcohol consumption to alcohol use disorder: Shared and unique genetic architectures in a cross-ancestry sample. American Journal of Psychiatry, 180(8), 584–593. doi:10.1176/appi.ajp.21090892

Kendler, K. S. (2022a). Incremental advances in psychiatric molecular genetics and nosology. World Psychiatry, 21(3), 415–416. doi:10.1002/wps.20999

Kendler, K. S. (2022b). Potential lessons for DSM from contemporary philosophy of science. JAMA Psychiatry, 79(2), 99–100. doi:10.1001/jamapsychiatry.2021.3559

Kotov, R., Krueger, R. F., Watson, D., Achenbach, T. M., Althoff, R. R., Bagby, R. M., … Zimmerman, M. (2017). The Hierarchical Taxonomy of Psychopathology (HiTOP): A dimensional alternative to traditional nosologies. Journal of Abnormal Psychology, 126(4), 454–477. doi:10.1037/abn0000258

Kraguljac, N. V., Guerreri, M., Strickland, M. J., & Zhang, H. (2023). Neurite orientation dispersion and density imaging in psychiatric disorders: A systematic literature review and a technical note. Biological Psychiatry Global Open Science, 3(1), 10–21. doi:10.1016/j.bpsgos.2021.12.012

La Manno, G., Gyllborg, D., Codeluppi, S., Nishimura, K., Salto, C., Zeisel, A., … Linnarsson, S. (2016). Molecular diversity of midbrain development in mouse, human, and stem cells. Cell, 167(2), 566–580.e519. doi:10.1016/j.cell.2016.09.027

Lam, M., Chen, C.-Y., Hill, W. D., Xia, C., Tian, R., Levey, D. F., … Lencz, T. (2022). Collective genomic segments with differential pleiotropic patterns between cognitive dimensions and psychopathology. Nature Communications, 13(1), 6868. doi:10.1038/s41467-022-34418-y

Lawrence, J. M., Foote, I. F., Breunig, S., Schaffer, L. S., Mallard, T. T., & Grotzinger, A. D. (2024). Shared genetic liability across systems of psychiatric and physical illness. medRxiv, 2024.2008.2002.24311427. doi:10.1101/2024.08.02.24311427

Levey, D. F., Galimberti, M., Deak, J. D., Wendt, F. R., Bhattacharya, A., Koller, D., … Gelernter, J. (2023). Multi-ancestry genome-wide association study of cannabis use disorder yields insight into disease biology and public health implications. Nature Genetics, 55(12), 2094–2103. doi:10.1038/s41588-023-01563-z

Li, M., Santpere, G., Imamura Kawasawa, Y., Evgrafov, O. V., Gulden, F. O., Pochareddy, S., … Li, Z. (2018). Integrative functional genomic analysis of human brain development and neuropsychiatric risks. Science, 362(6420), eaat7615. doi:10.1126/science.aat7615

Liang, Y., Melia, O., Caroll, T. J., Brettin, T., Brown, A., & Im, H. K. (2022). BrainXcan identifies brain features associated with behavioral and psychiatric traits using large scale genetic and imaging data. medRxiv, 2021.2006.2001.21258159. doi:10.1101/2021.06.01.21258159

Liu, Z., Liu, R., Gao, H., Jung, S., Gao, X., Sun, R., … Chinese Inflammatory Bowel Disease Genetics, C. (2023). Genetic architecture of the inflammatory bowel diseases across East Asian and European ancestries. Nature Genetics, 55(5), 796–806. doi:10.1038/s41588-023-01384-0

Mahajan, A., Spracklen, C. N., Zhang, W., Ng, M. C. Y., Petty, L. E., Kitajima, H., … Morris, A. P. (2022). Multi-ancestry genetic study of type 2 diabetes highlights the power of diverse populations for discovery and translation. Nature Genetics, 54(5), 560–572. doi:10.1038/s41588-022-01058-3

Meng, X., Navoly, G., Giannakopoulou, O., Levey, D. F., Koller, D., Pathak, G. A., … Kuchenbaecker, K. (2024). Multi-ancestry genome-wide association study of major depression aids locus discovery, fine mapping, gene prioritization and causal inference. Nature Genetics, 56(2), 222–233. doi:10.1038/s41588-023-01596-4

Michelini, G., Palumbo, I. M., DeYoung, C. G., Latzman, R. D., & Kotov, R. (2021). Linking RDoC and HiTOP: A new interface for advancing psychiatric nosology and neuroscience. Clinical Psychology Review, 86, 102025. doi:10.1016/j.cpr.2021.102025

Mulugeta, A., Zhou, A., King, C., & Hyppönen, E. (2020). Association between major depressive disorder and multiple disease outcomes: a phenome-wide Mendelian randomisation study in the UK Biobank. Molecular Psychiatry, 25(7), 1469–1476. doi:10.1038/s41380-019-0486-1

Picci, G., Taylor, B. K., Killanin, A. D., Eastman, J. A., Frenzel, M. R., Wang, Y.-P., … Wilson, T. W. (2022). Left amygdala structure mediates longitudinal associations between exposure to threat and long-term psychiatric symptomatology in youth. Human Brain Mapping, 43(13), 4091–4102. doi:10.1002/hbm.25904

Sambuco, N., Mickle, A. M., Garvan, C., Cardoso, J., Johnson, A. J., Kusko, D. A., … Sibille, K. T. (2022). Vulnerable dispositional traits and chronic pain: Predisposing but not predetermining. The Journal of Pain, 23(4), 693–705. doi:10.1016/j.jpain.2021.11.007

Sanchez-Roige, S., Jennings, M. V., Thorpe, H. H. A., Mallari, J. E., van der Werf, L. C., Bianchi, S. B., … andMe Research, T. (2023). CADM2 is implicated in impulsive personality and numerous other traits by genome- and phenome-wide association studies in humans and mice. Translational Psychiatry, 13(1), 167. doi:10.1038/s41398-023-02453-y

The GTEx Consortium, Aguet, F., Anand, S., Ardlie, K. G., Gabriel, S., Getz, G. A., … Volpi, S. (2020). The GTEx Consortium atlas of genetic regulatory effects across human tissues. Science, 369(6509), 1318–1330. doi:10.1126/science.aaz1776

Toikumo, S., Vickers-Smith, R., Jinwala, Z., Xu, H., Saini, D., Hartwell, E., … Kranzler, H. R. (2024). A multi-ancestry genetic study of pain intensity in 598,339 veterans. Nature Medicine, 30(7), 1075– 1084. doi:10.1038/s41591-024-02839-5

Trubetskoy, V., Pardiñas, A. F., Qi, T., Panagiotaropoulou, G., Awasthi, S., Bigdeli, T. B., … Bertolino, A. (2022). Mapping genomic loci implicates genes and synaptic biology in schizophrenia. Nature, 604(7906), 502–508. doi:10.1038/s41586-022-04434-5

Tully, P. J., Harrison, N. J., Cheung, P., & Cosh, S. (2016). Anxiety and cardiovascular disease risk: A review. Current Cardiology Reports, 18(12), 120. doi:10.1007/s11886-016-0800-3

Waszczuk, M. A., Miao, J., Docherty, A. R., Shabalin, A. A., Jonas, K. G., Michelini, G., & Kotov, R. (2023). General v. specific vulnerabilities: Polygenic risk scores and higher-order psychopathology dimensions in the Adolescent Brain Cognitive Development (ABCD) Study. Psychological Medicine, 53(5), 1937–1946. doi:10.1017/S0033291721003639

Watanabe, K., Umićević Mirkov, M., de Leeuw, C. A., van den Heuvel, M. P., & Posthuma, D. (2019). Genetic mapping of cell type specificity for complex traits. Nature Communications, 10(1), 3222. doi:10.1038/s41467-019-11181-1

Wendt, F. R., Pathak, G. A., Lencz, T., Krystal, J. H., Gelernter, J., & Polimanti, R. (2021). Multivariate genome-wide analysis of education, socioeconomic status and brain phenome. Nature Human Behaviour, 5(4), 482–496. doi:10.1038/s41562-020-00980-y

Williams, C. M., Labouret, G., Wolfram, T., Peyre, H., & Ramus, F. (2023). A general cognitive ability factor for the UK Biobank. Behavior Genetics, 53(2), 85–100. doi:10.1007/s10519-022-10127-6

Williams, F. M. K., Elgaeva, E. E., Freidin, M. B., Zaytseva, O. O., Aulchenko, Y. S., Tsepilov, Y. A., & Suri, P. (2022). Causal effects of psychosocial factors on chronic back pain: a bidirectional Mendelian randomisation study. European Spine Journal, 31(7), 1906–1915. doi:10.1007/s00586-022-07263-2

Yao, C., Zhang, Y., Lu, P., Xiao, B., Sun, P., Tao, J., … Fang, M. (2023). Exploring the bidirectional relationship between pain and mental disorders: A comprehensive Mendelian randomization study. The Journal of Headache and Pain, 24(1), 82. doi:10.1186/s10194-023-01612-2

Zhang, F., Baranova, A., Zhou, C., Cao, H., Chen, J., Zhang, X., & Xu, M. (2021). Causal influences of neuroticism on mental health and cardiovascular disease. Human Genetics, 140(9), 1267–1281. doi:10.1007/s00439-021-02288-x

Zhou, W., Kanai, M., Wu, K. H., Rasheed, H., Tsuo, K., Hirbo, J. B., … Neale, B. M. (2022). Global Biobank Meta-analysis Initiative: Powering genetic discovery across human disease. Cell Genomics, 2(10), 100192. doi:10.1016/j.xgen.2022.100192

Zhu, Z., Zheng, Z., Zhang, F., Wu, Y., Trzaskowski, M., Maier, R., … Yang, J. (2018). Causal associations between risk factors and common diseases inferred from GWAS summary data. Nature Communications, 9(1), 224. doi:10.1038/s41467-017-02317-2

