## Supplementary Figures for "Integrating HiTOP and RDoC Frameworks Part II: Shared and Distinct Biological Mechanisms of Externalizing and Internalizing Psychopathology"

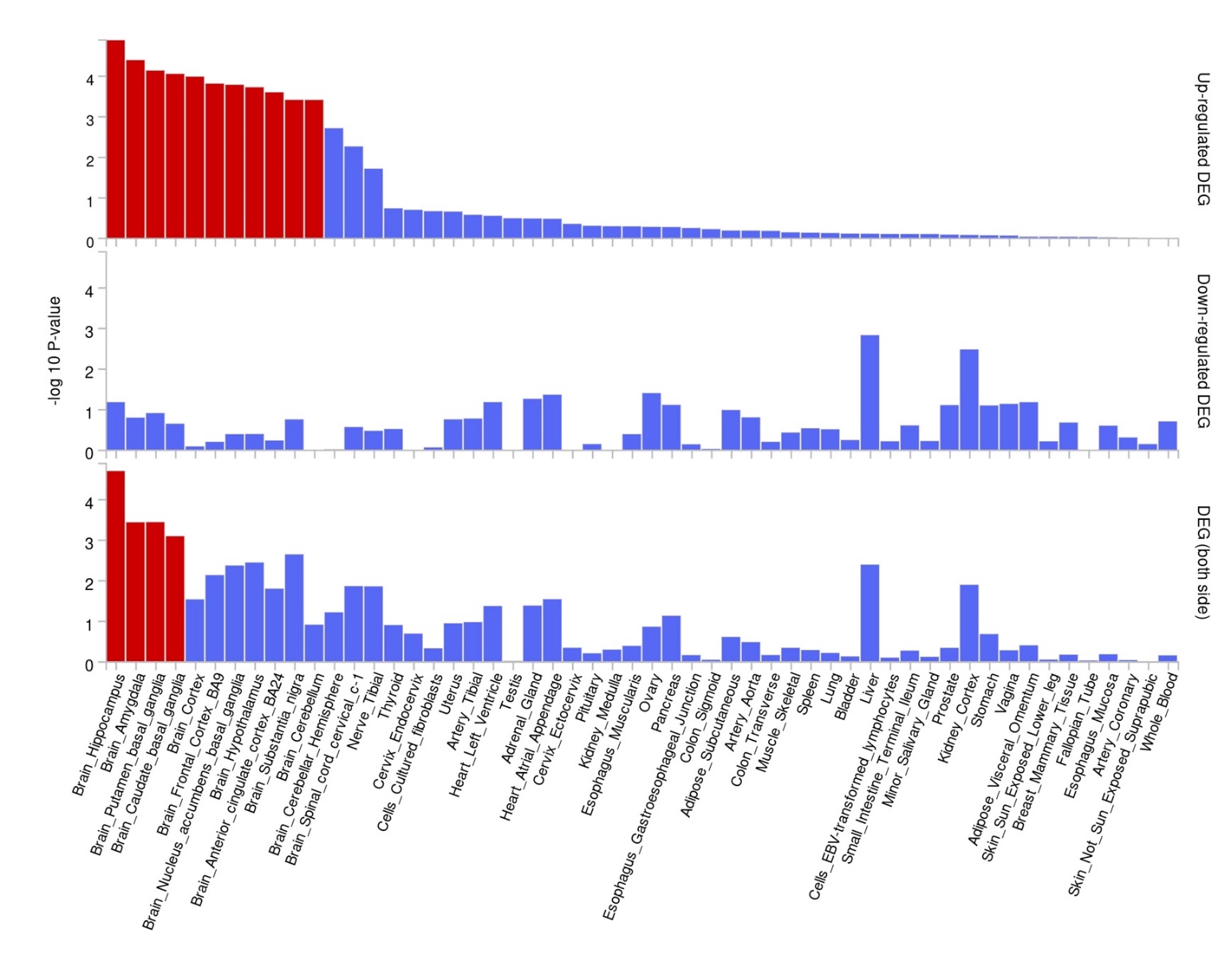

### **Supplementary Figure 1. Results of differential gene expression across tissue types for externalizing.**

Red indicates significance at a Bonferroni-adjusted p-value of 9.26e-04.

a)

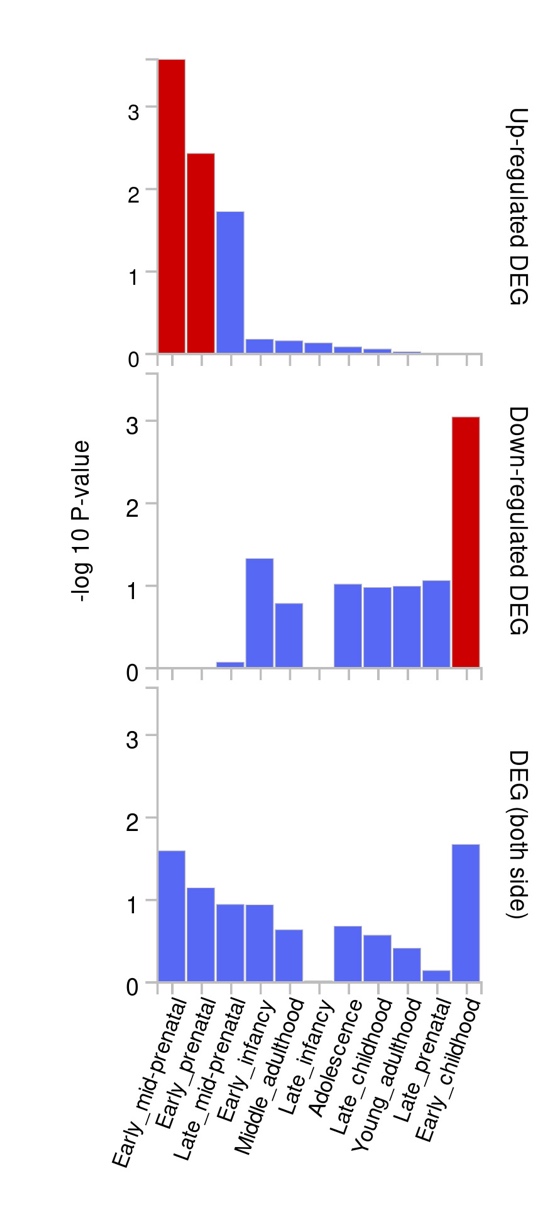

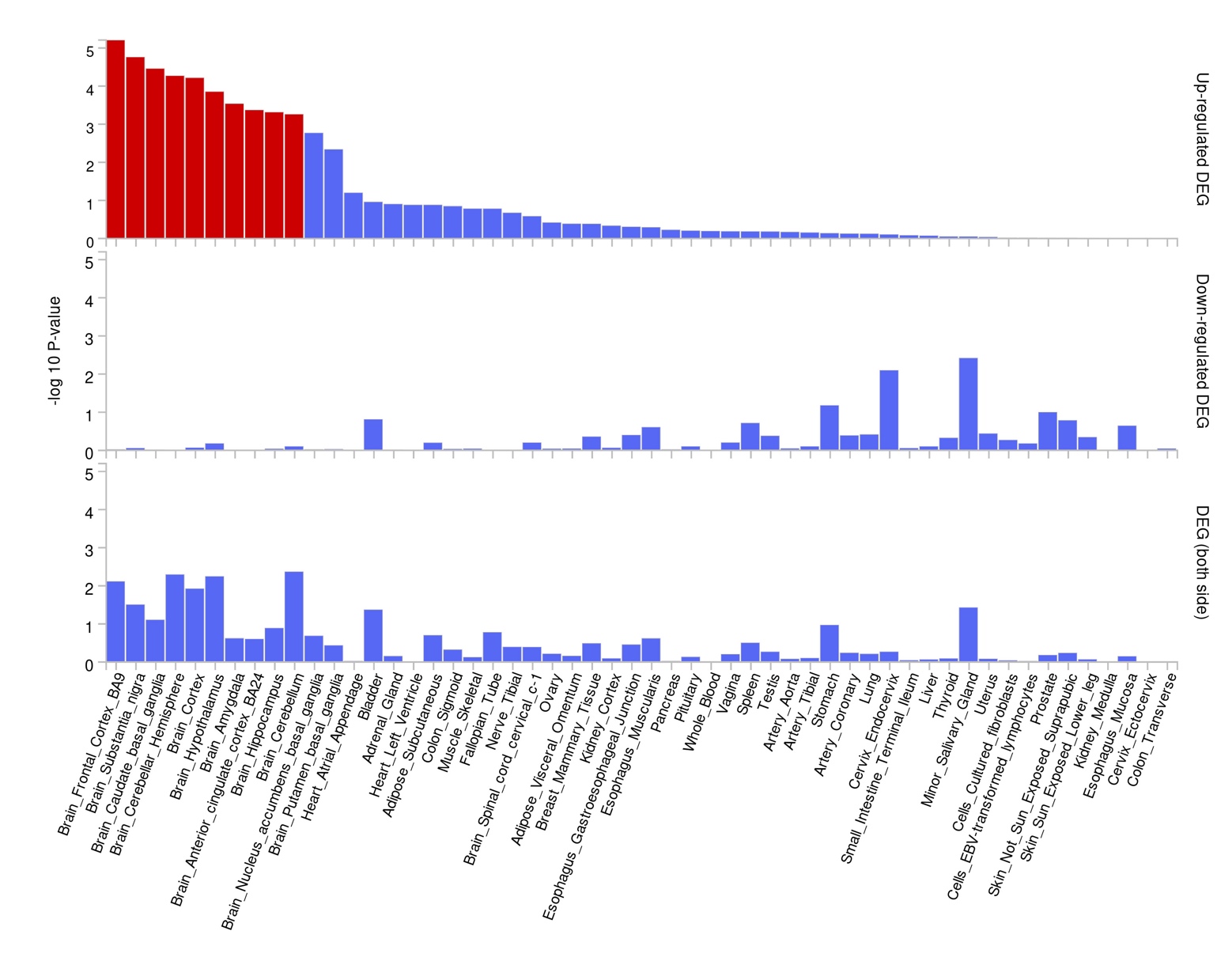

b)

### **Supplementary Figure 2*.* Differential gene expression across tissue types (a) and developmental stages (b) for externalizing and internalizing (EXT+INT).**

Red indicates significance at a Bonferroni-adjusted p-value of 4.55e-03 for developmental stages and 9.26e-04 for tissue types.

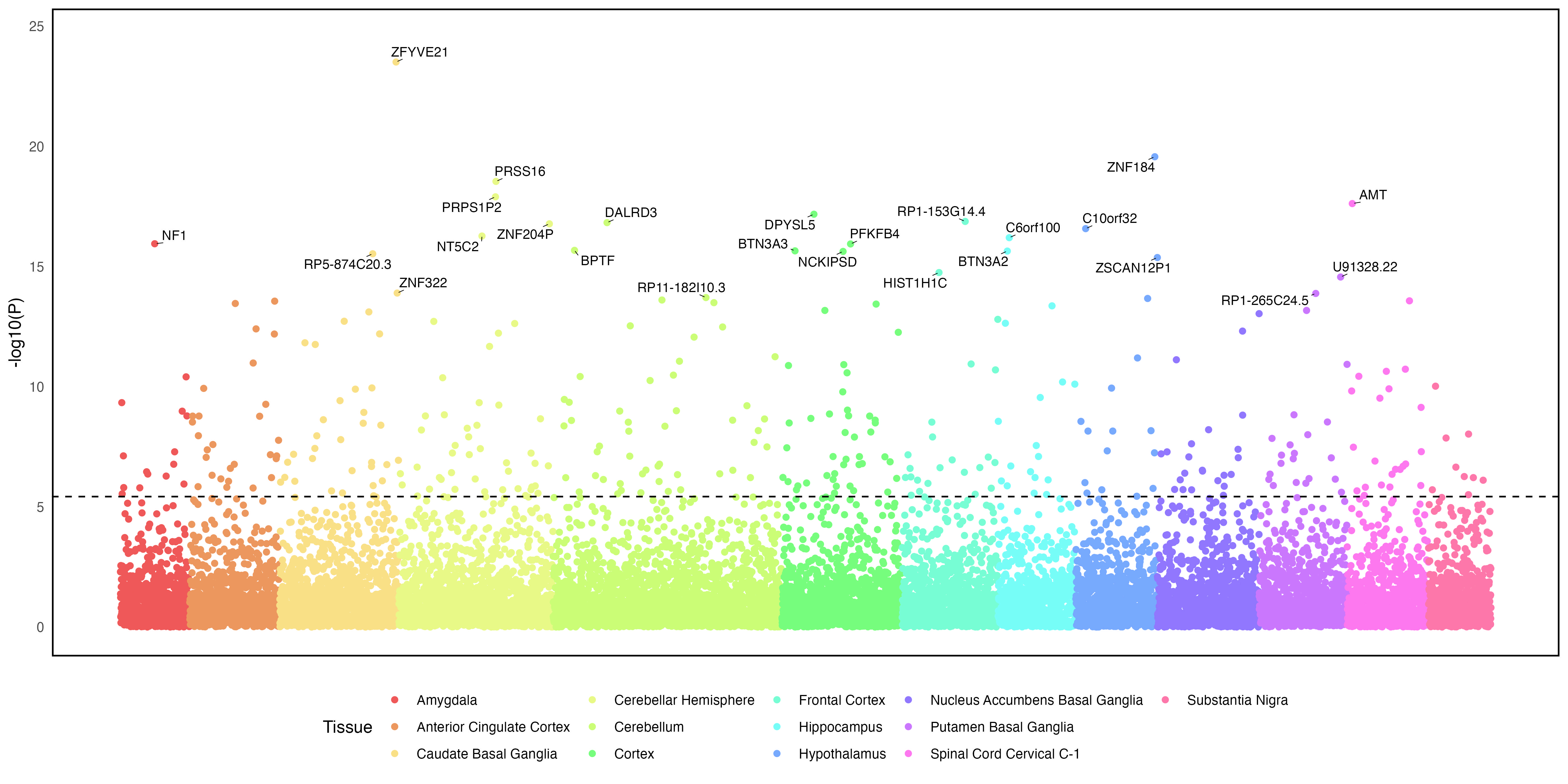

a)

b)

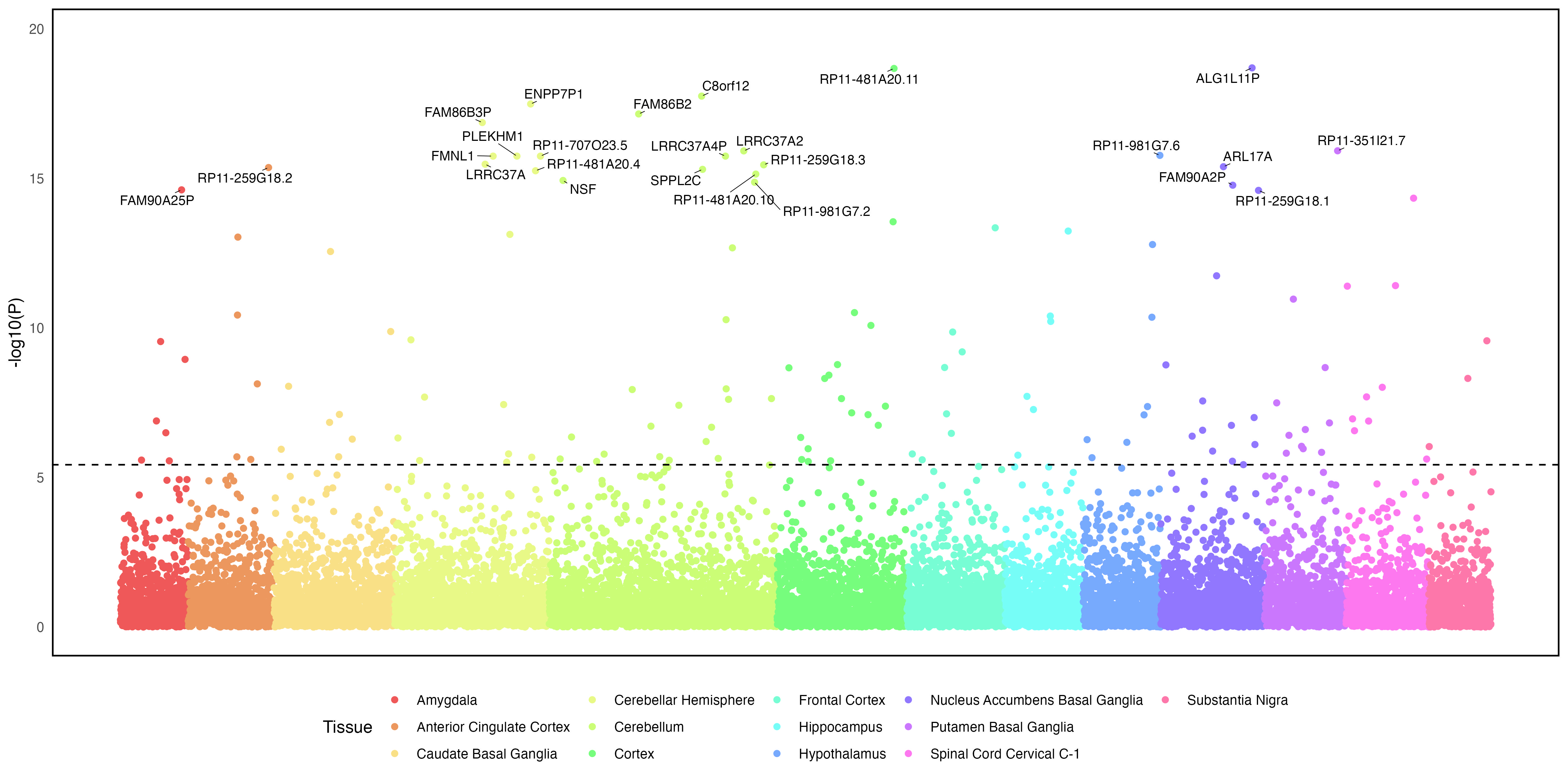

### **Supplementary Figure 3. Transcriptome wide association analysis using S-MultiXcan for externalizing (Panel A) and internalizing (Panel B).**

Analyses examined effects on gene expression across 13 brain tissues. The top 25 associations are annotated for each factor. The best associated tissue type for each gene is indicated by color.

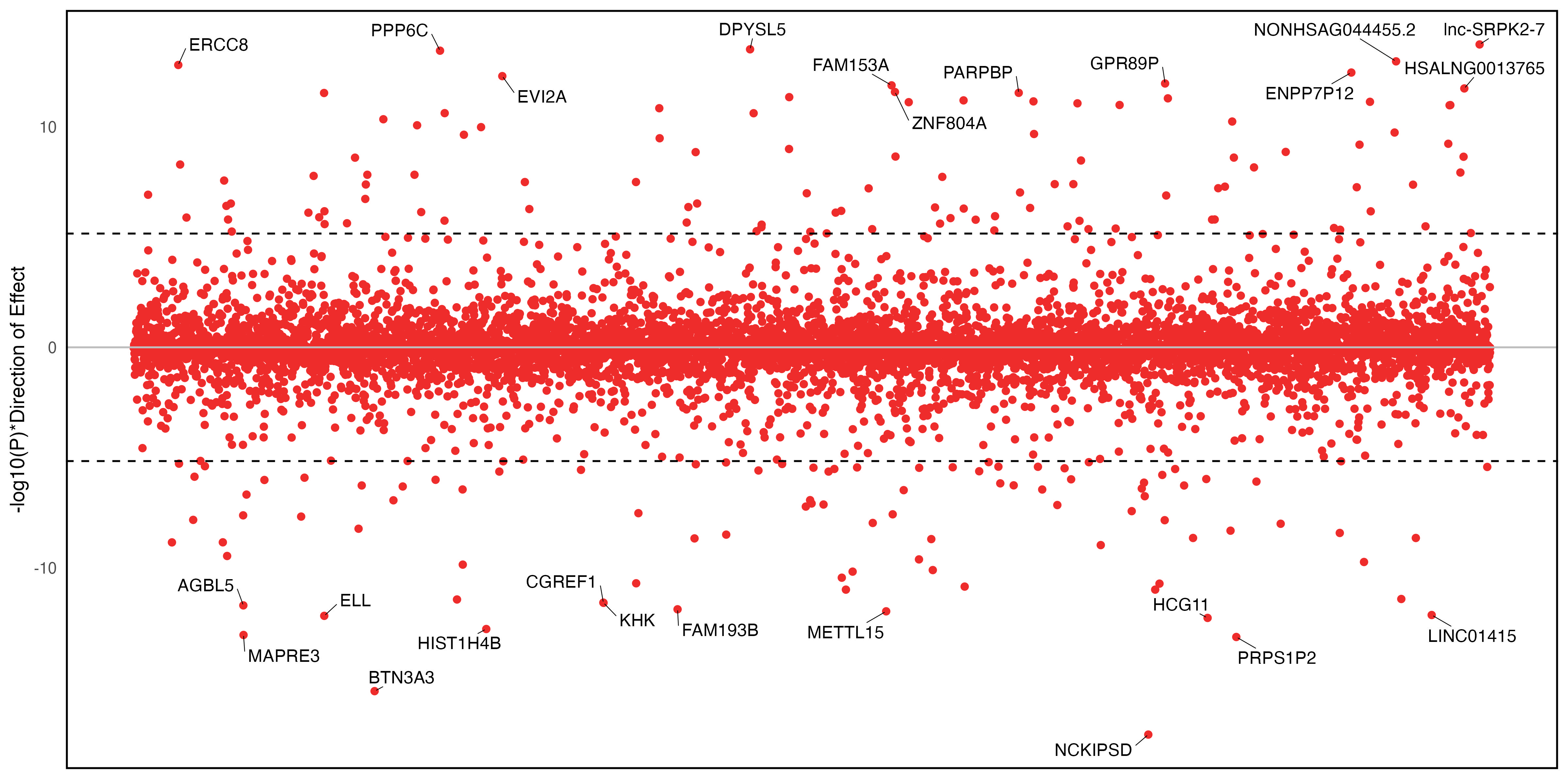

b)

a)

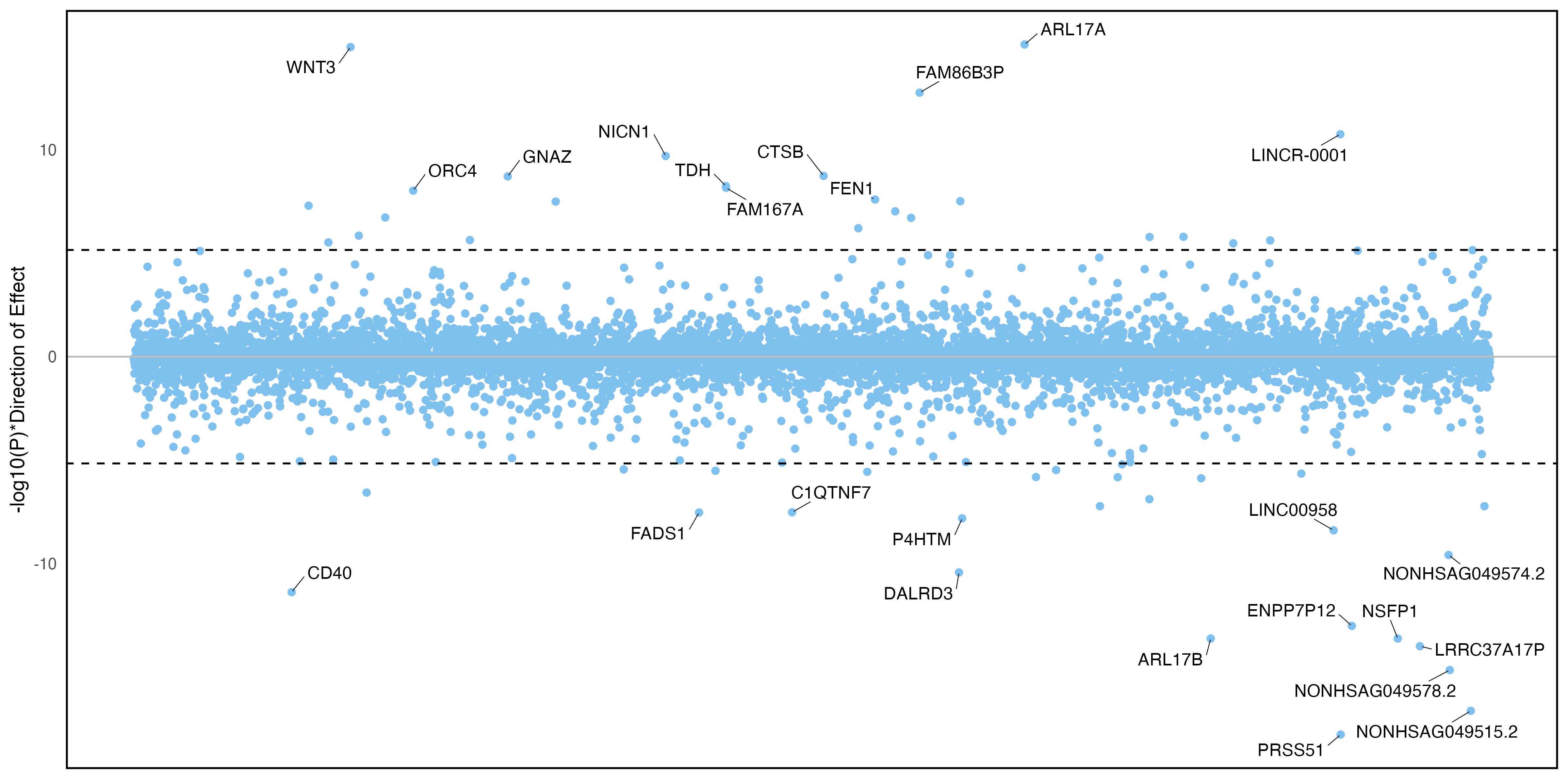

c)

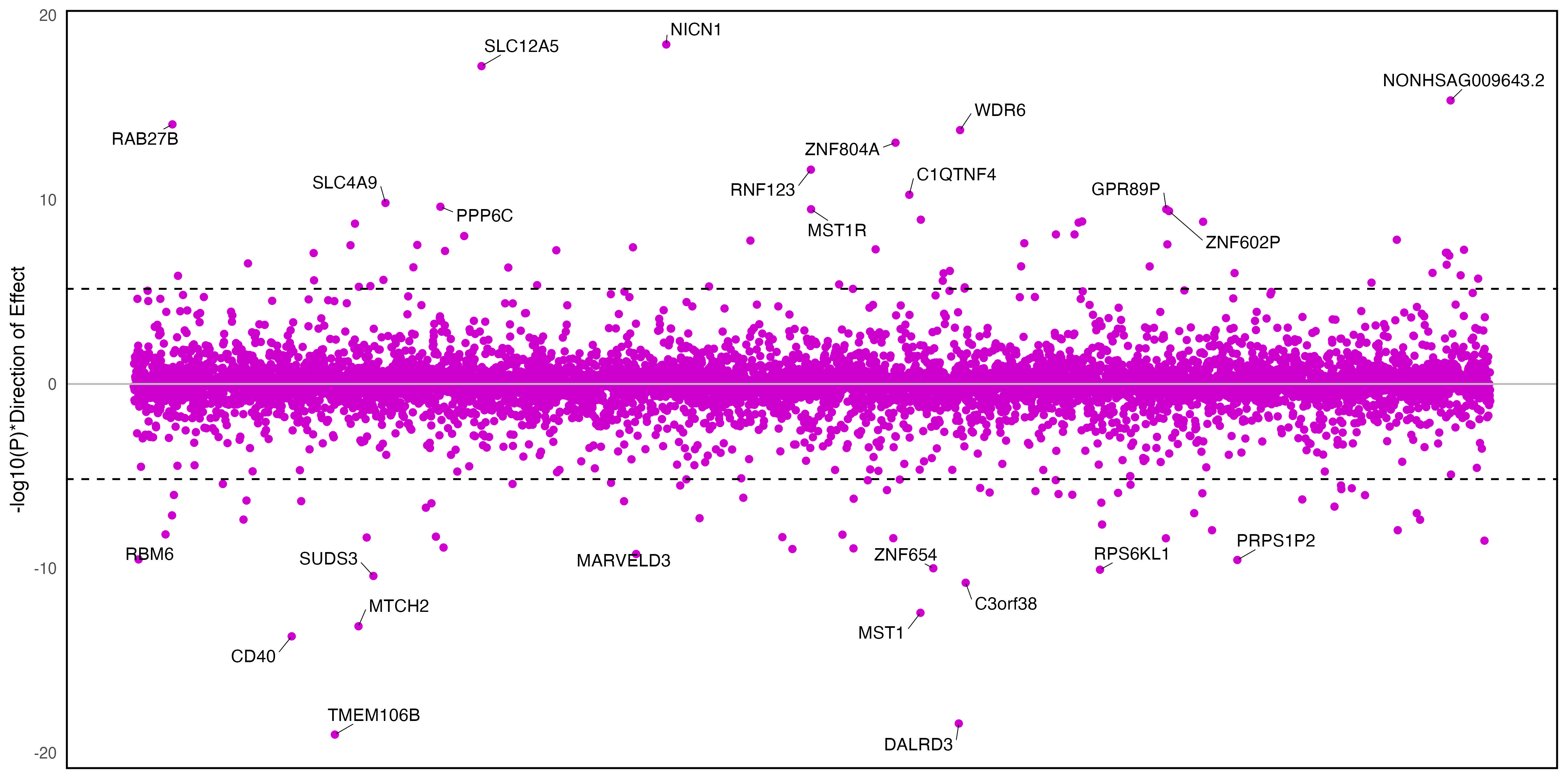

### **Supplementary Figure 4. Transcriptome wide association analysis using data from psychiatric cases and controls for S-PrediXcan.**

Y-axis plots the p-value and whether genes were up- or down-regulated in brain tissue. The top 25 associations are annotated. Panel A is externalizing, B is internalizing, and C is EXT+INT.

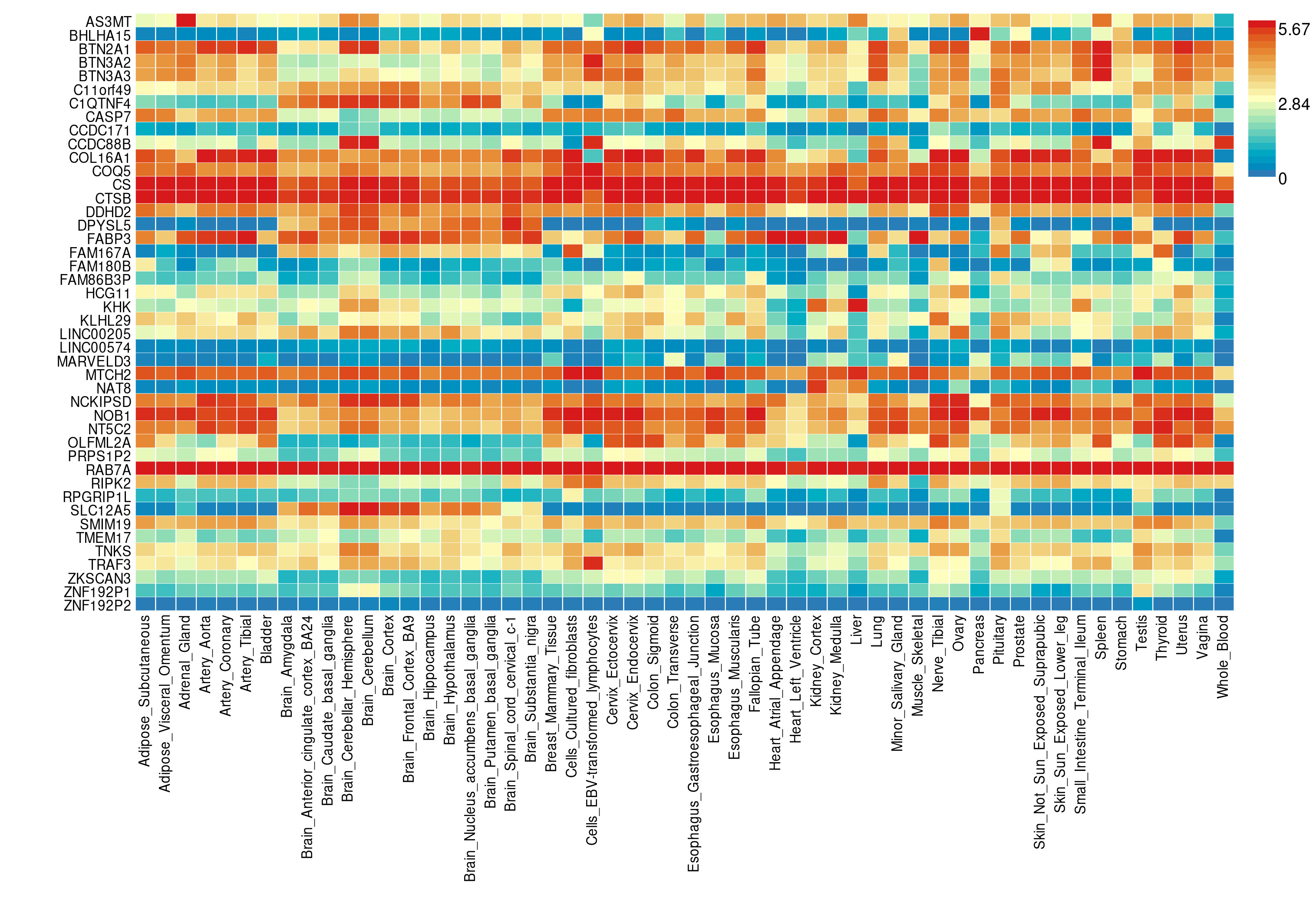

### **Supplementary Figure 5. Gene-property analyses performed on the subset of genes that were identified by both TWAS for externalizing.**

Heatmap presents average expression levels (log^2^ transformed).

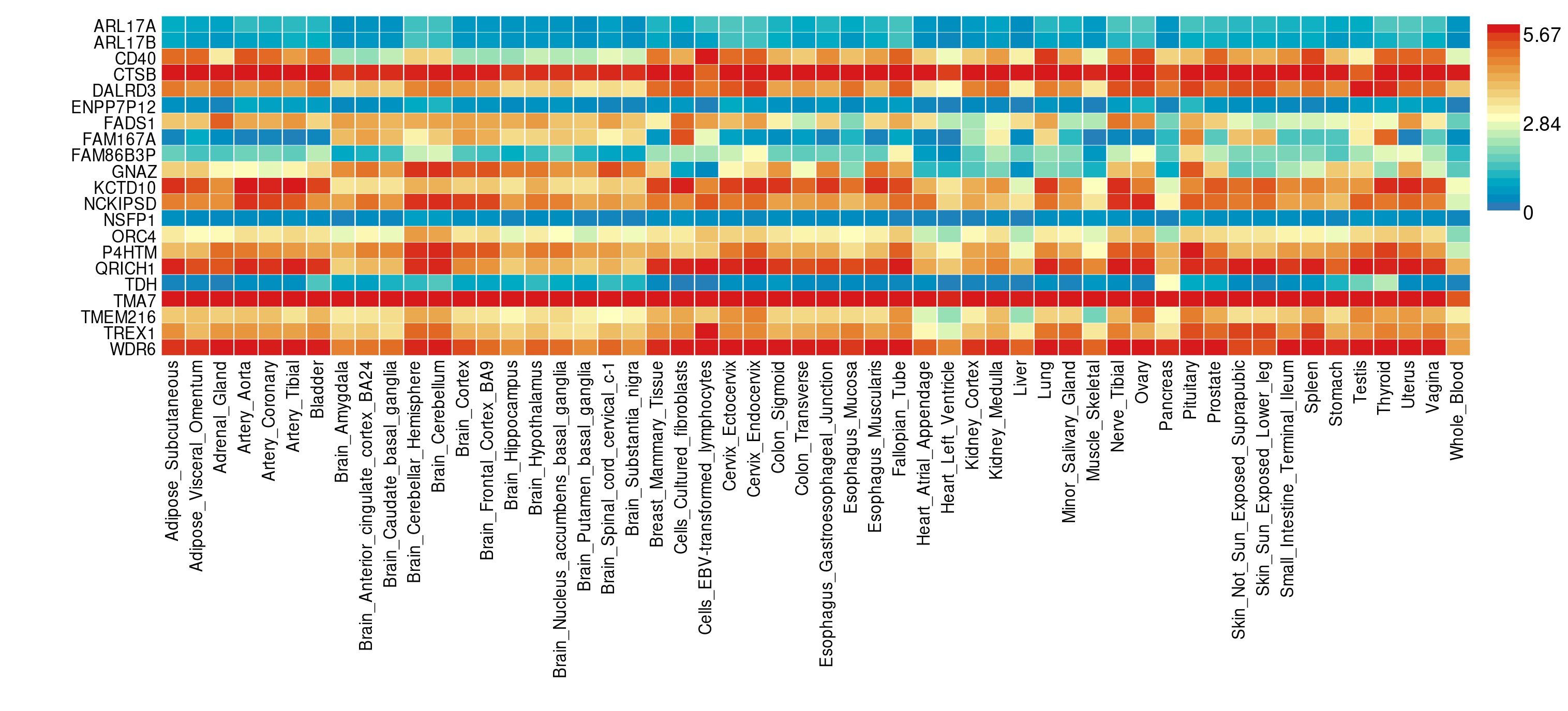

### **Supplementary Figure 6. Gene-property analyses performed on the subset of genes that were identified by both TWAS for internalizing.**

Heatmap presents average expression levels (log^2^ transformed).

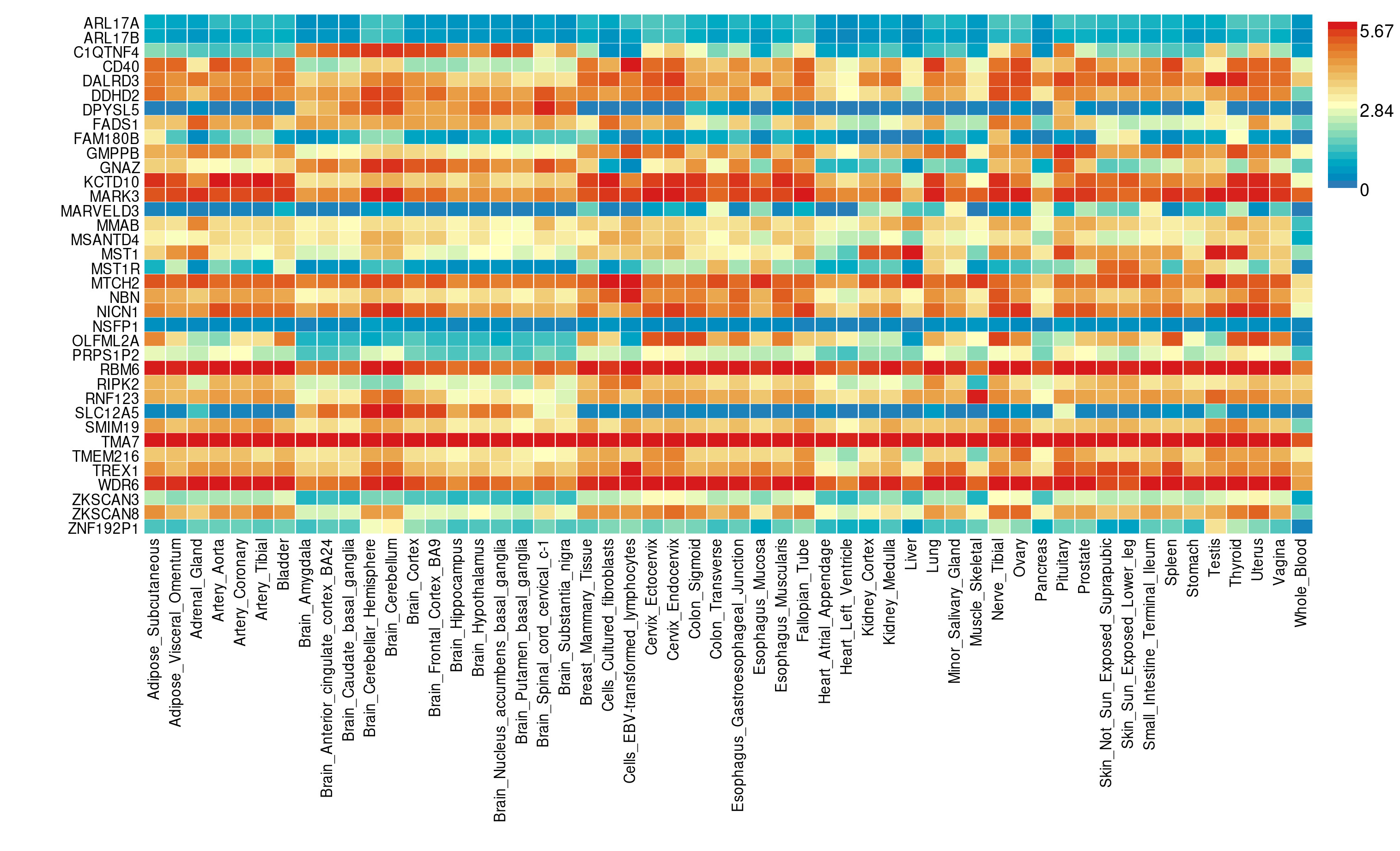

### **Supplementary Figure 7. Gene-property analyses performed on the subset of genes that were identified by both TWAS for EXT+INT.**

Heatmap presents average expression levels (log^2^ transformed).

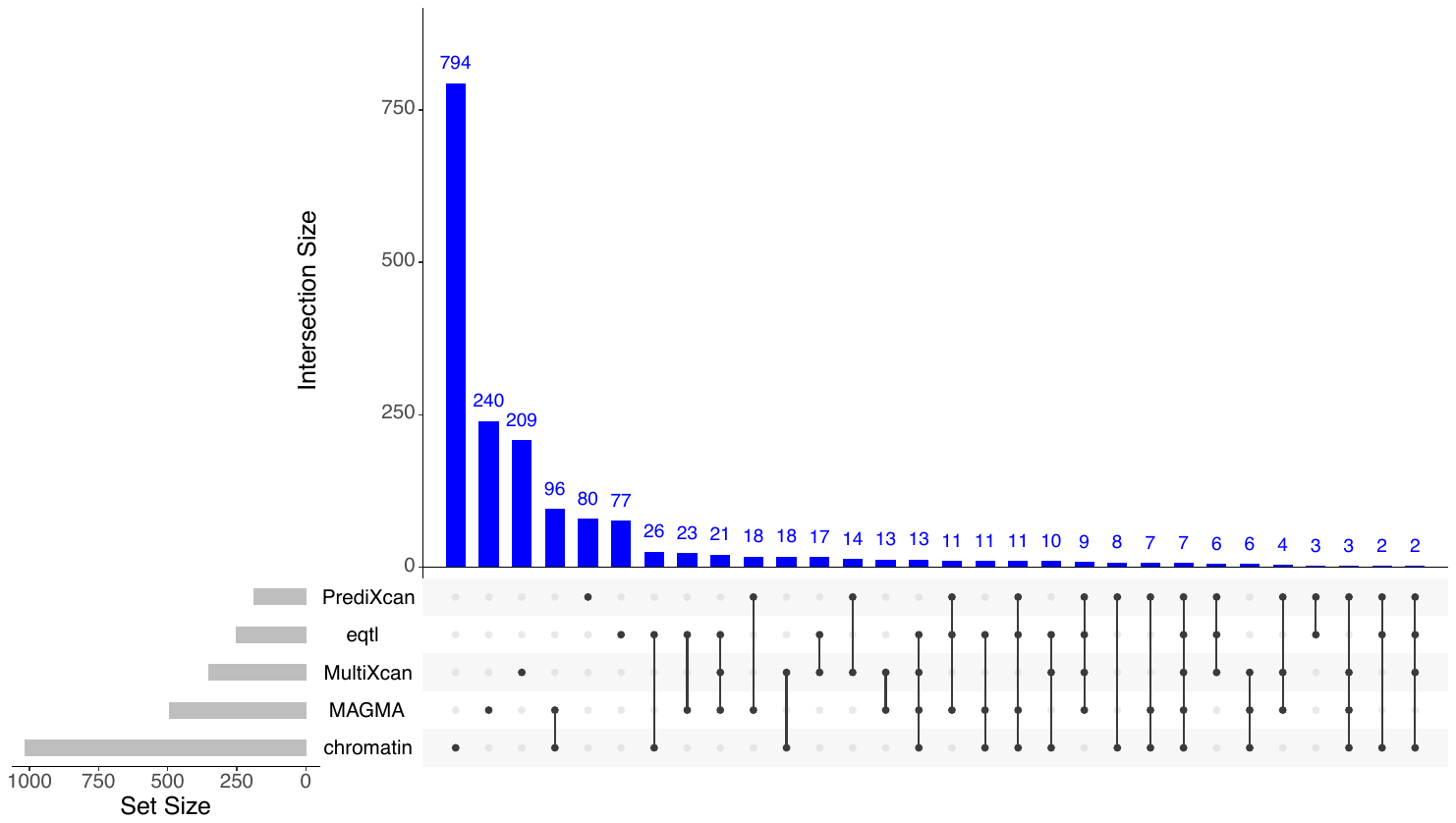

Externalizing

Internalizing

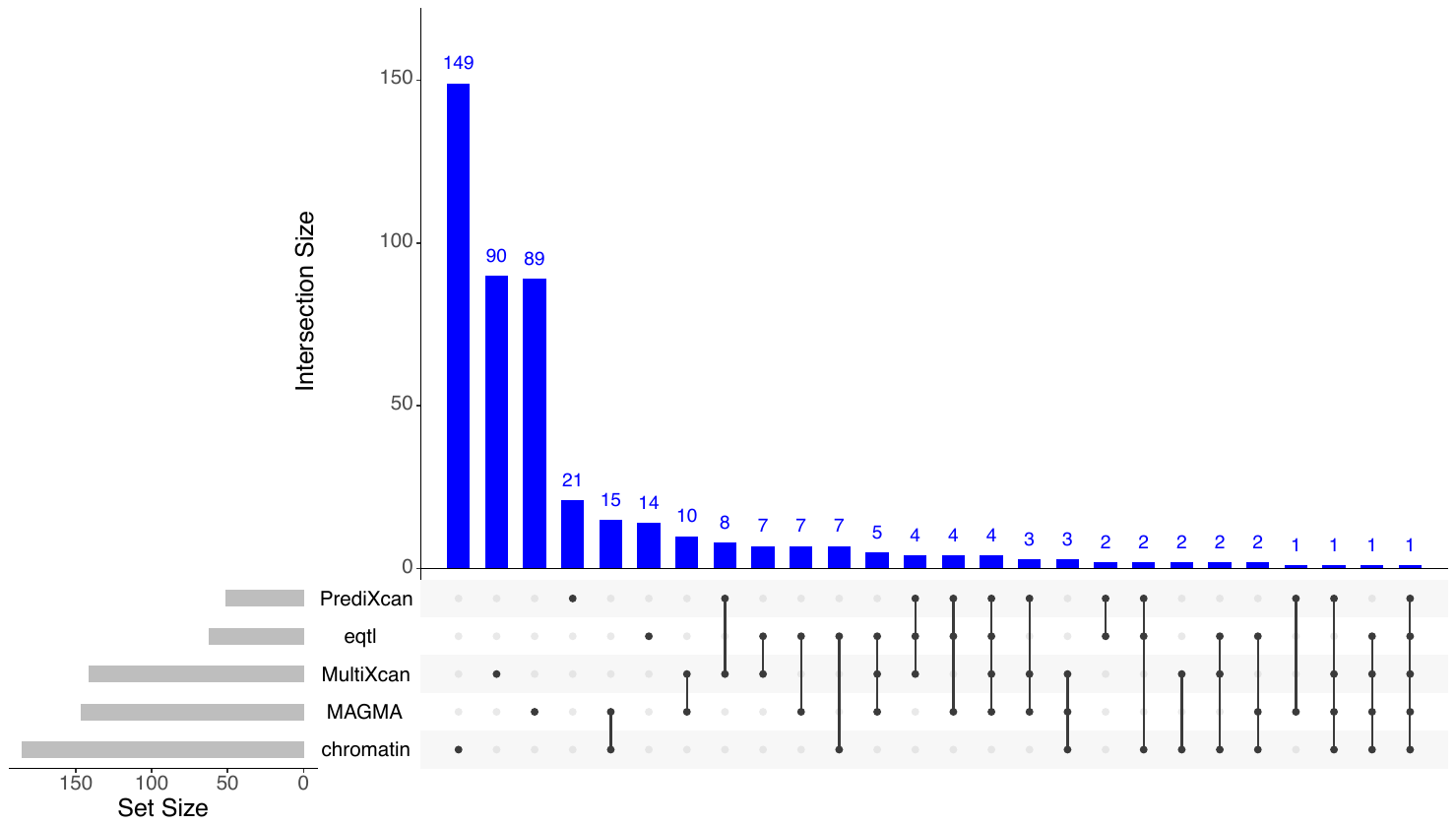

### **Supplementary Figure 8. UpSet plots of genes identified for externalizing (EXT) and internalizing (INT).**

PrediXcan and MultiXcan genes were identified in the transcriptome wide association studies conducted using brain tissue data from psychiatric cases/controls and generally healthy individuals, respectively. eQTL genes were identified using expression quantitative trait loci mapping. MAGMA protein-coding genes were identified using a genome-wide significance gene-based test. Chromatin genes were identified using chromatin interaction mapping. Set size indicates the number of genes identified by each approach, and the interaction size indicates the number of genes identified by the set of approaches, noted by solid dots.

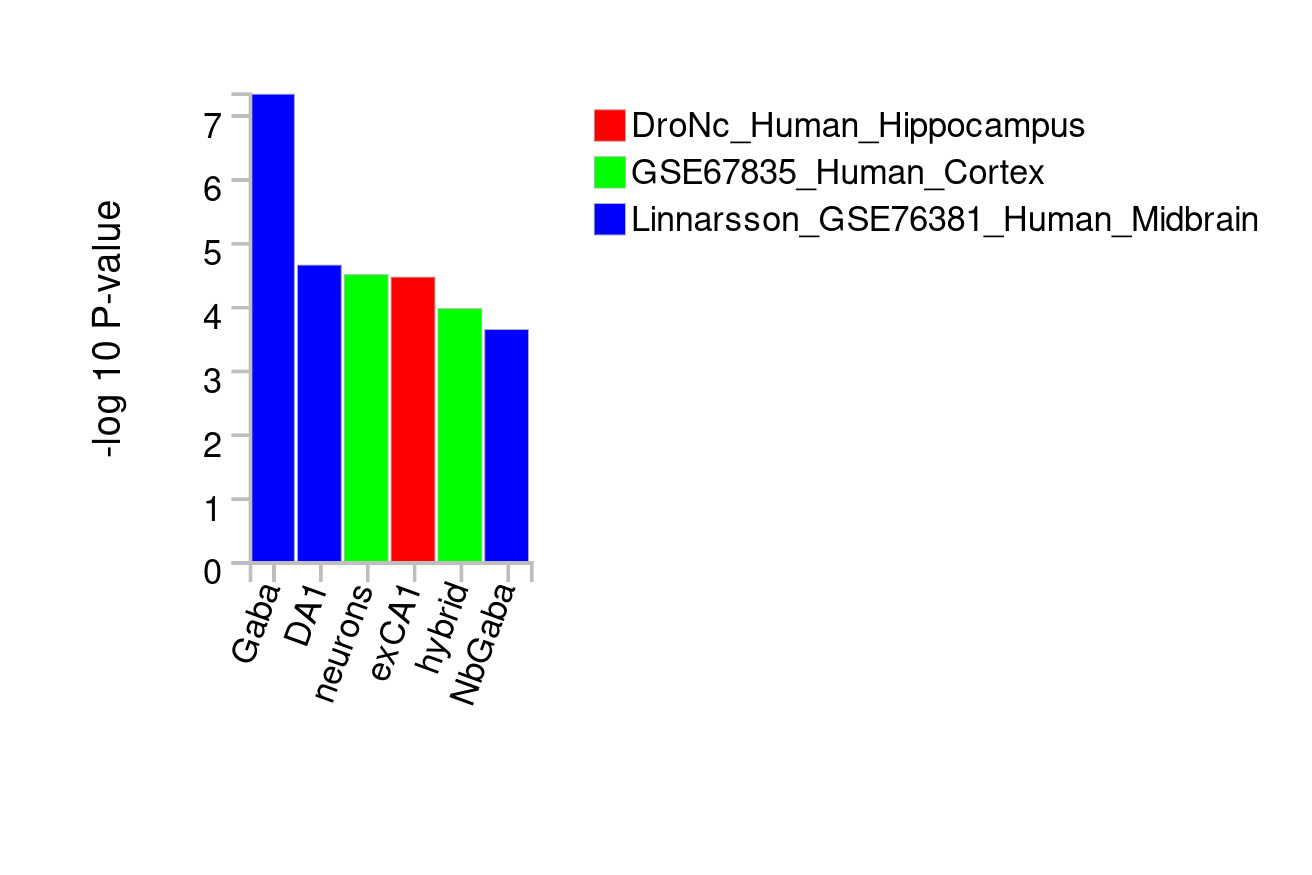

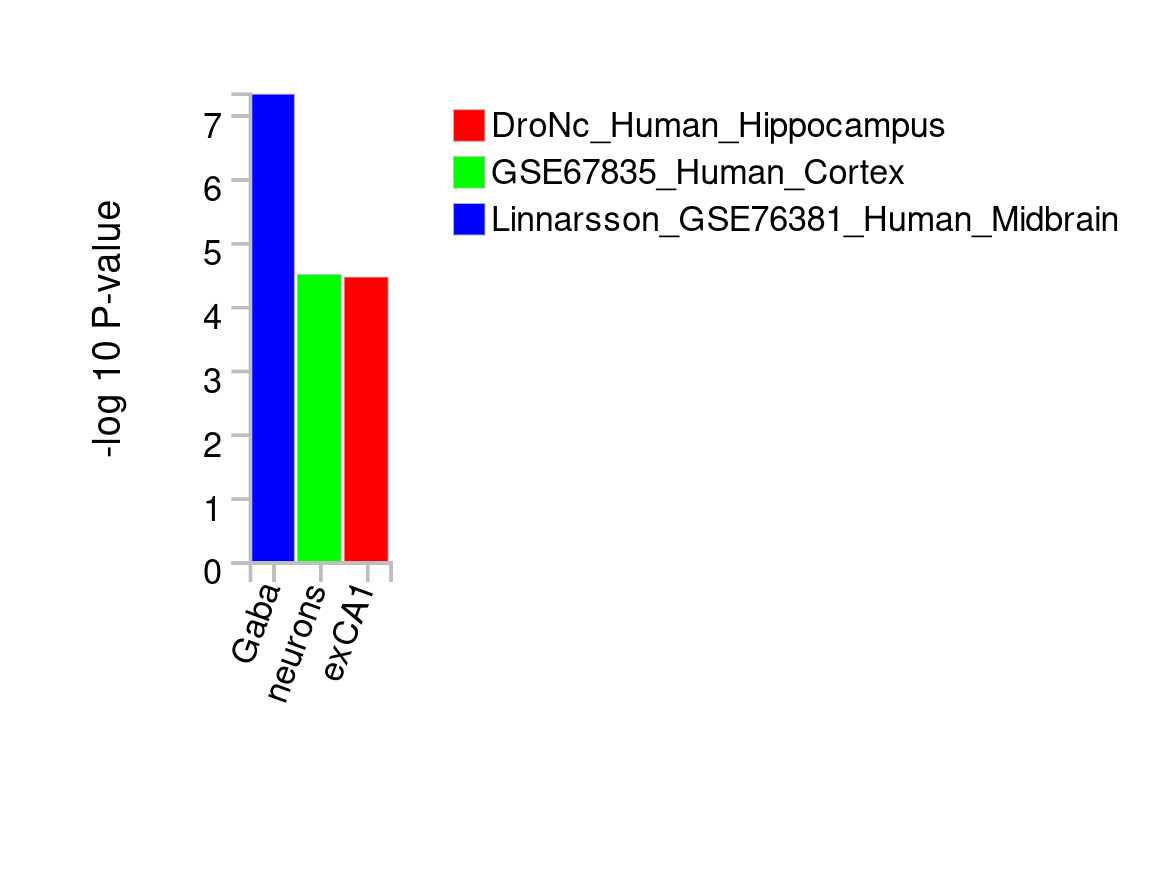

b)

a)

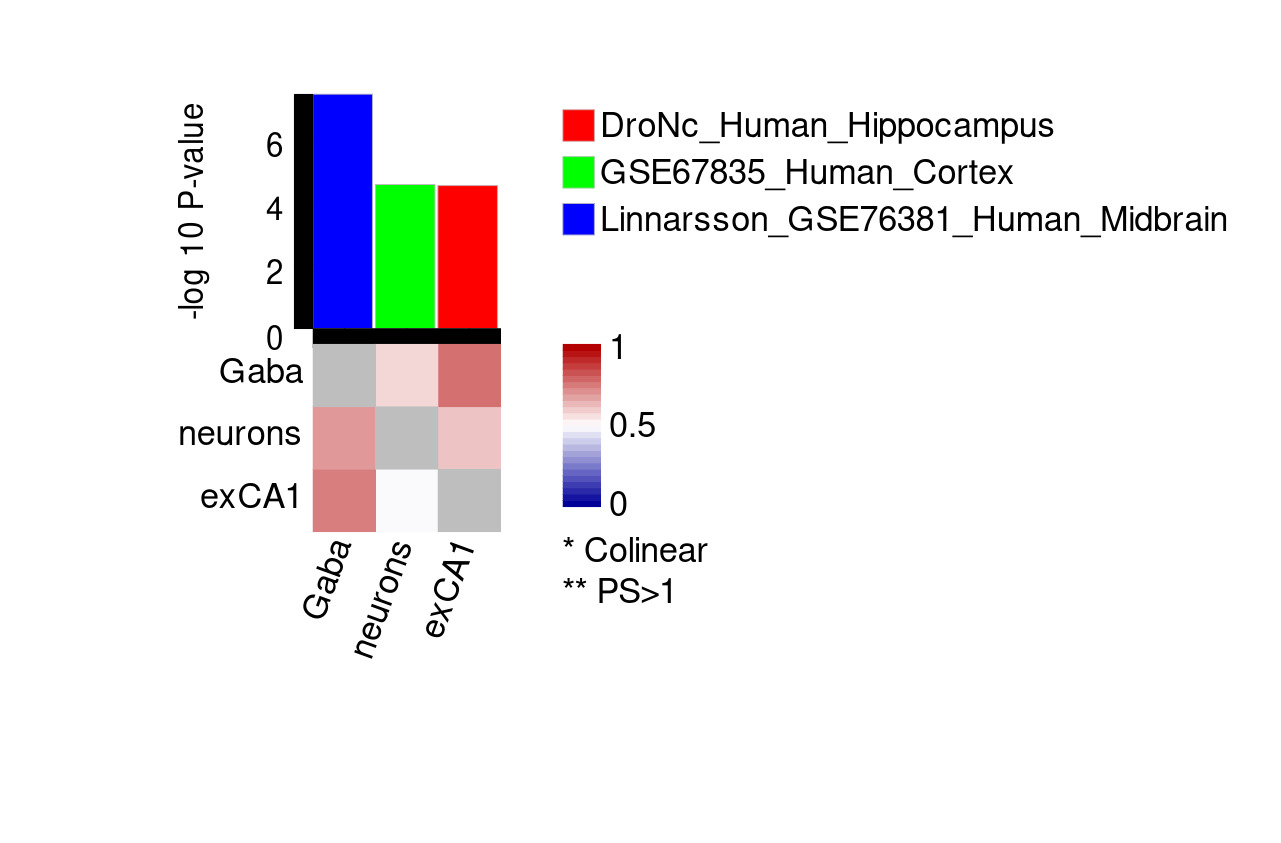

c)

### **Supplementary Figure 9. Results of cell-type specificity gene expression analyses for externalizing.**

Analyses used single cell RNA sequencing (scRNA-seq) datasets from human brain cells. Panel A shows results from step one across dataset analyses, panel B shows step 2 within-dataset conditional analyses, and panel C shows step 3 cross datasets conditional analyses. In panel C, the heatmap shows the cross-datasets proportional significance (PS) of cell type j (column) conditioning on cell type i (row). Each cell is colored by PS with upper limit 1, where PS>1 is represented by double stars. A star represents a pair of collinear cell types. Panel C bar plot presents marginal P-value of the cell type.

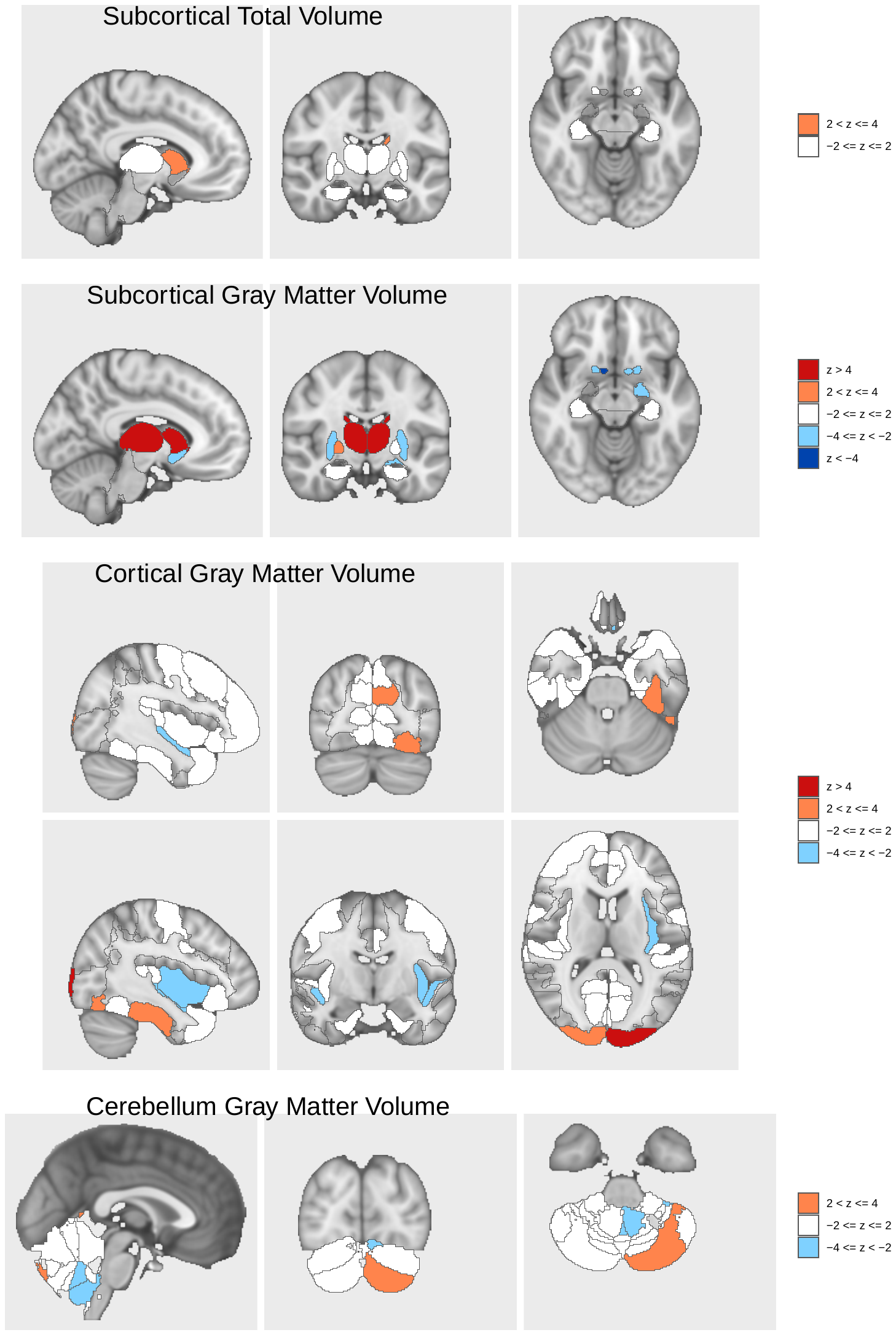

### **Supplementary Figure 10. BrainXcan structural MRI association results for externalizing.**

The direction and magnitude of the Z-scores of the association tests for each region are plotted by color. Interactive files showing the regions in the visualization are available [here](https://liangyy.github.io/brainxcan-docs/docs/example.html#63_Visualization_and_report).

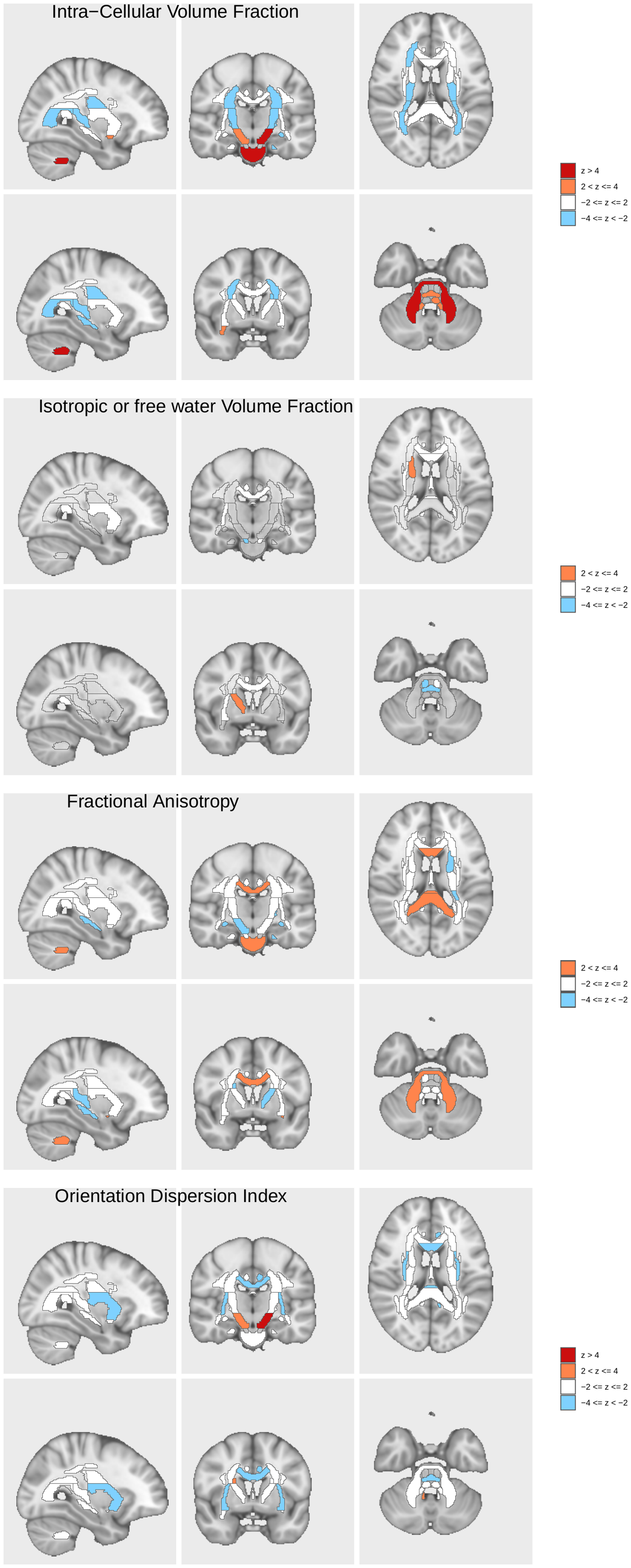

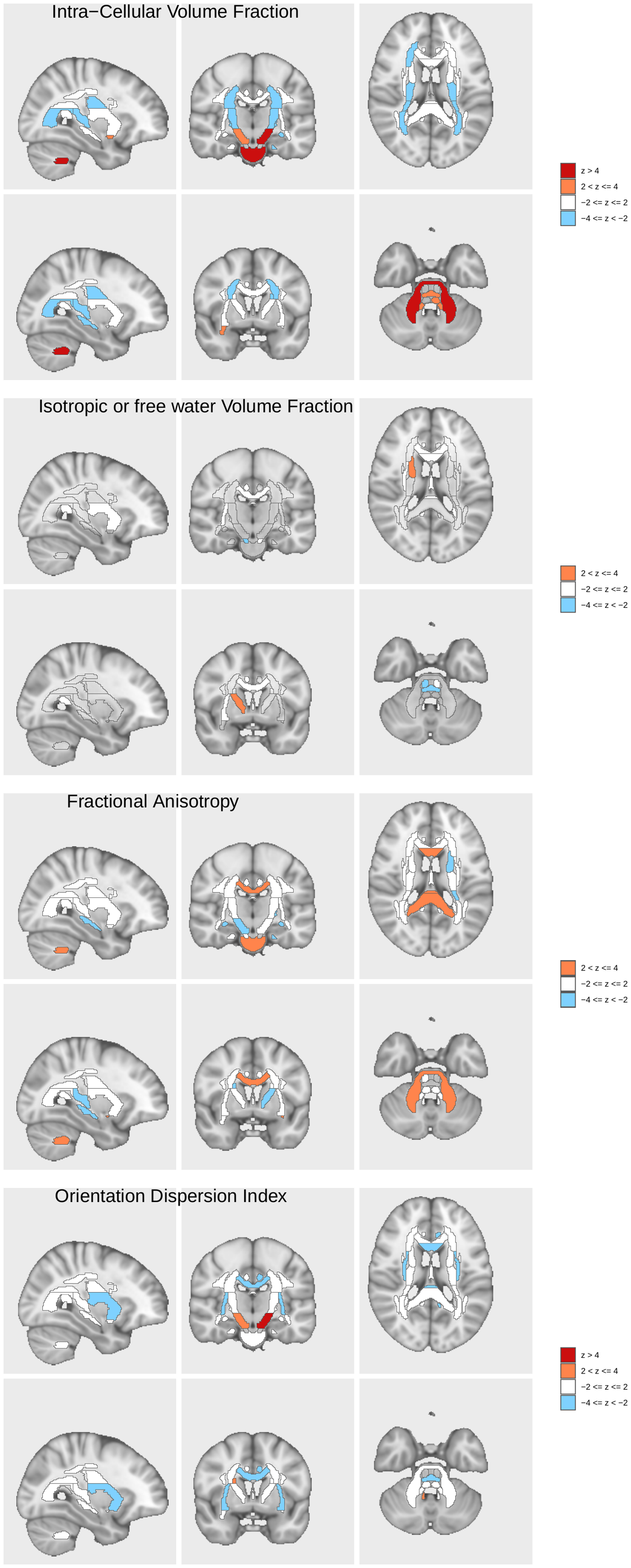

### **Supplementary Figure 11. BrainXcan diffusion MRI association results for externalizing.**

The direction and magnitude of the Z-scores of the association tests for each region are plotted by color. Interactive files showing the regions in the visualization are available [here](https://liangyy.github.io/brainxcan-docs/docs/example.html#63_Visualization_and_report).

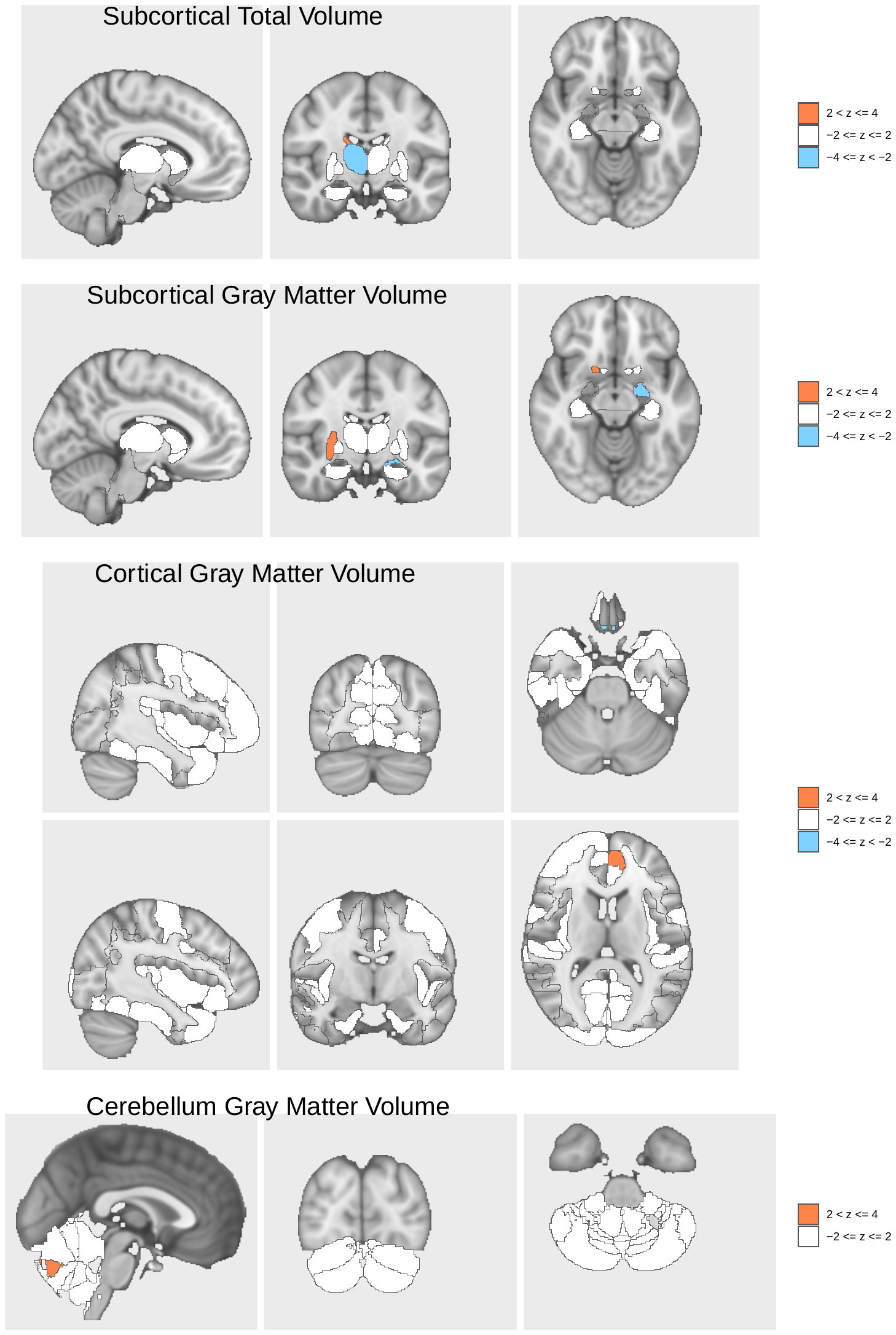

### **Supplementary Figure 12. BrainXcan structural MRI association results for internalizing.**

The direction and magnitude of the Z-scores of the association tests for each region are plotted by color. Interactive files showing the regions in the visualization are available [here](https://liangyy.github.io/brainxcan-docs/docs/example.html#63_Visualization_and_report).

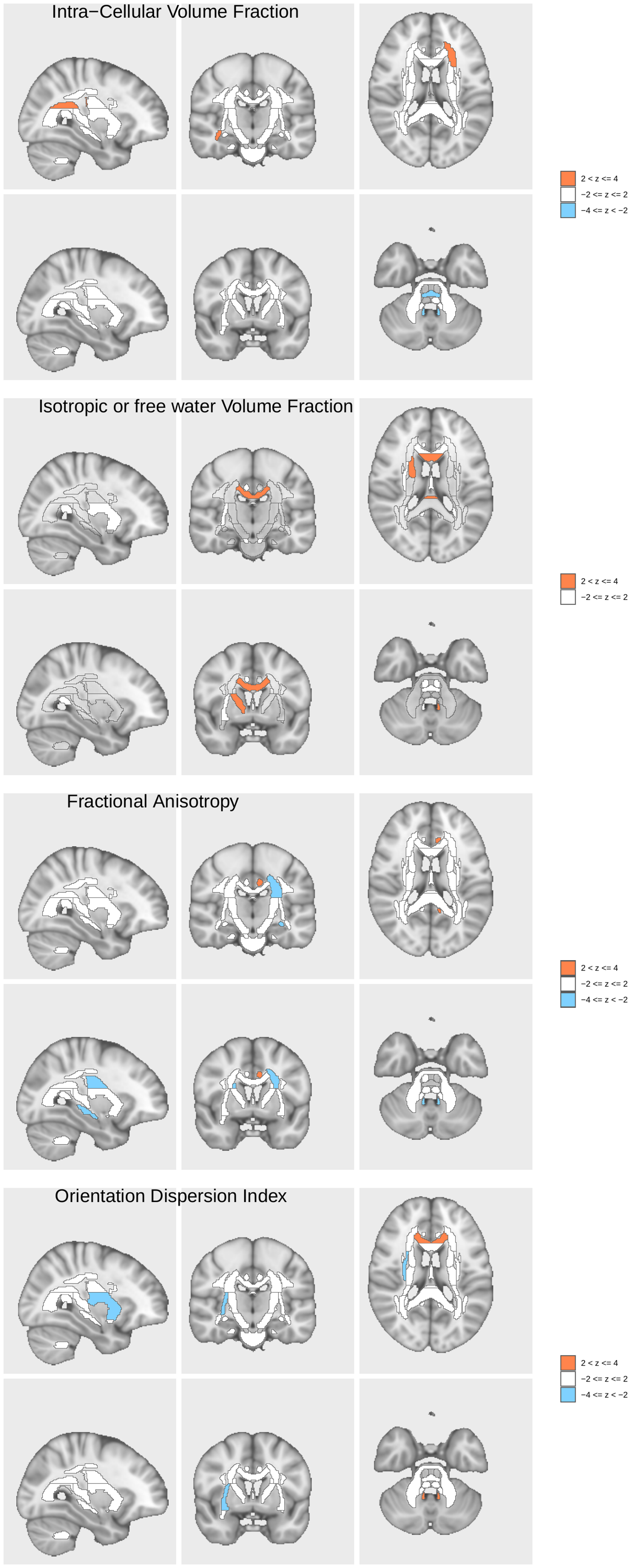

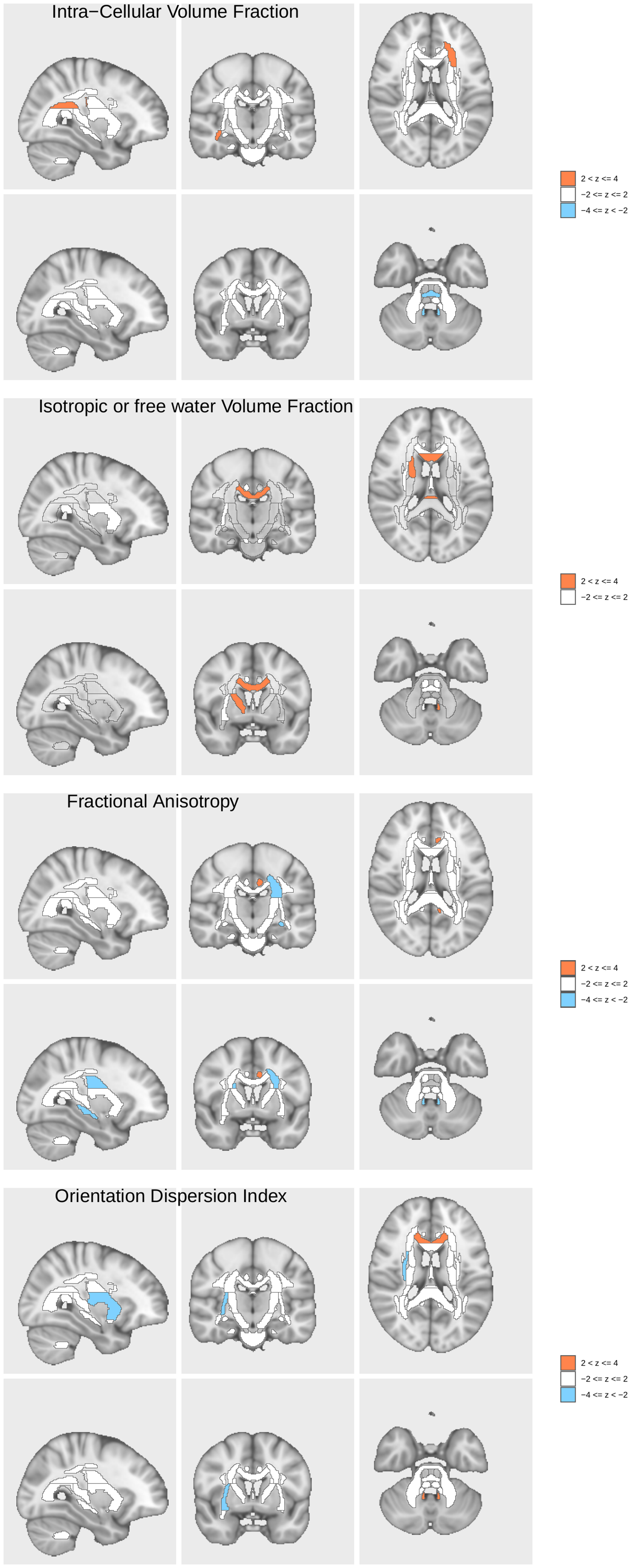

### **Supplementary Figure 13. BrainXcan diffusion MRI association results for internalizing.**

The direction and magnitude of the Z-scores of the association tests for each region are plotted by color. Interactive files showing the regions in the visualization are available [here](https://liangyy.github.io/brainxcan-docs/docs/example.html#63_Visualization_and_report).

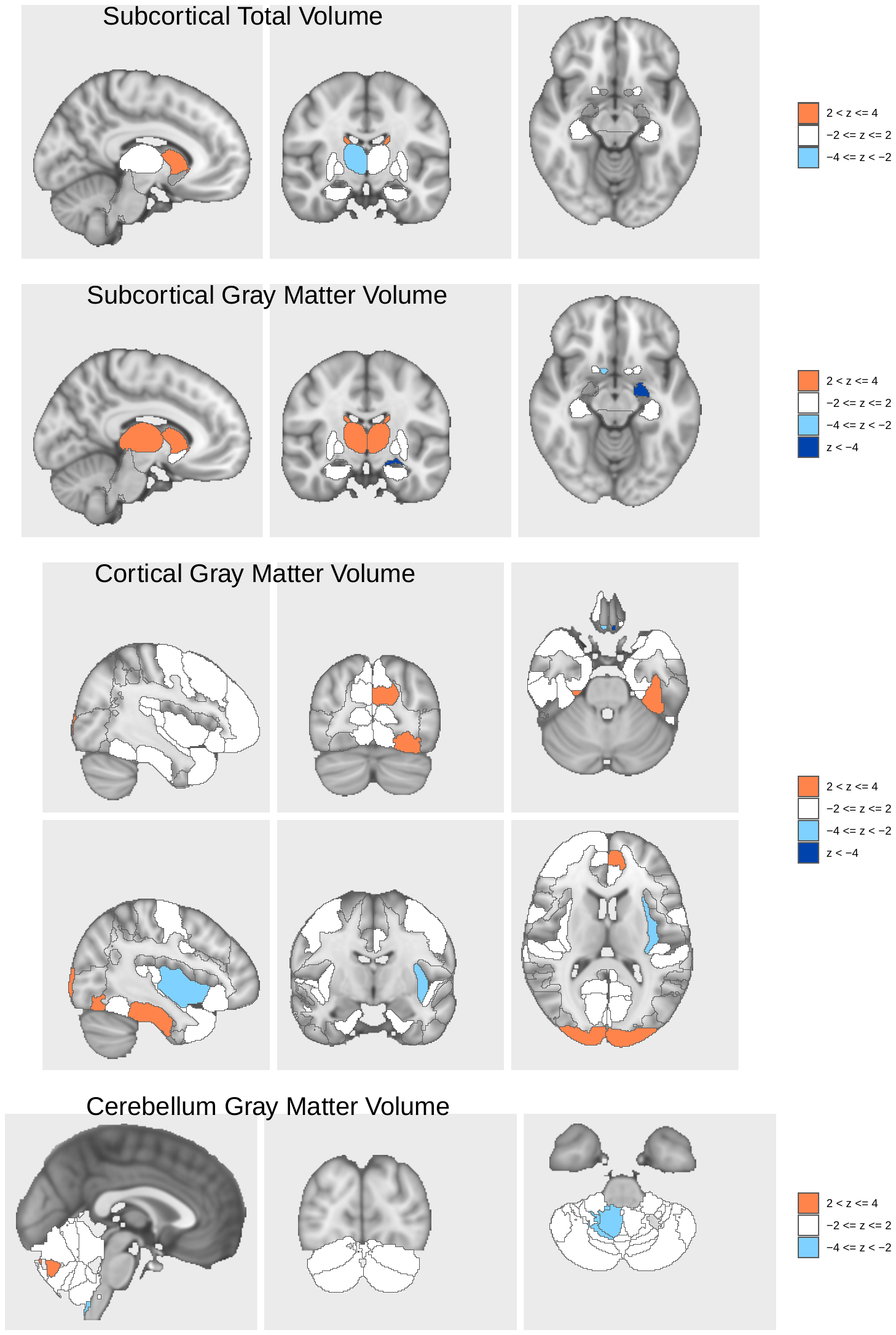

### **Supplementary Figure 14. BrainXcan structural MRI association results for second order externalizing + internalizing.**

The direction and magnitude of the Z-scores of the association tests for each region are plotted by color. Interactive files showing the regions in the visualization are available [here](https://liangyy.github.io/brainxcan-docs/docs/example.html#63_Visualization_and_report).

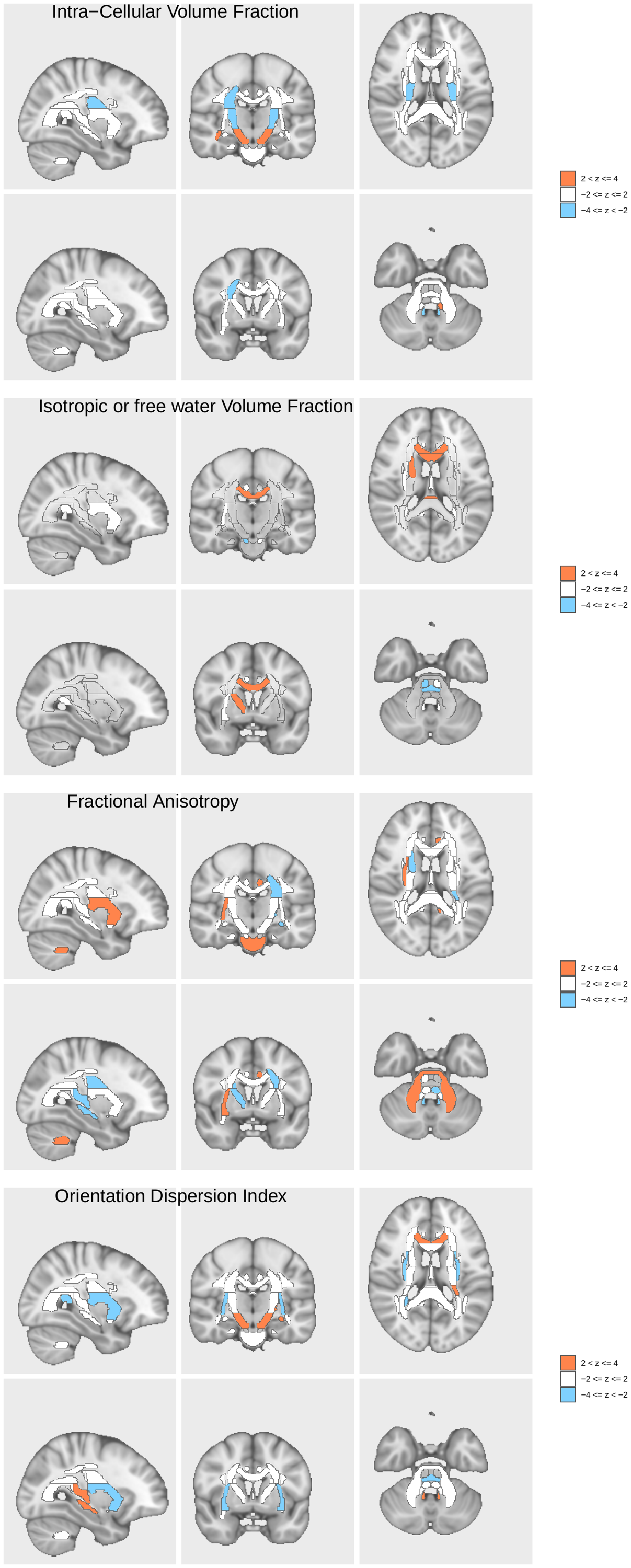

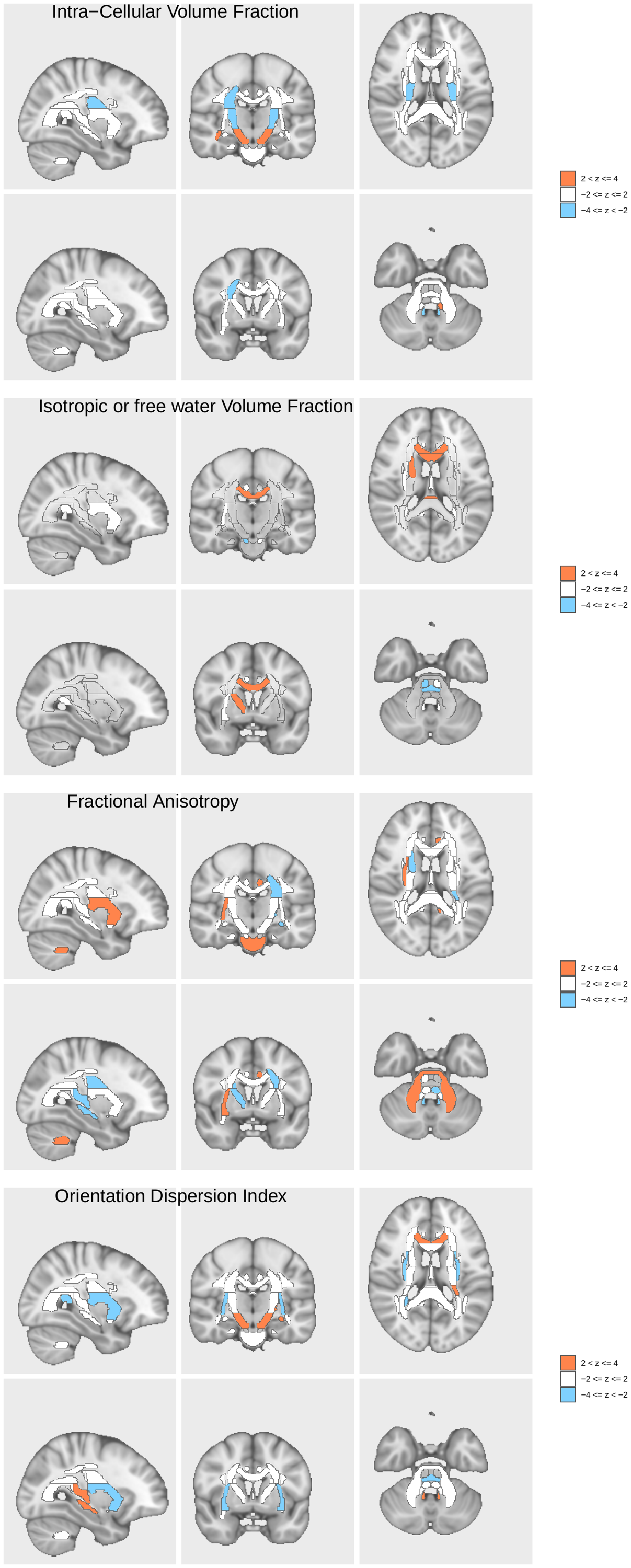

### **Supplementary Figure 15. BrainXcan diffusion MRI association results for second order externalizing + internalizing.**

The direction and magnitude of the Z-scores of the association tests for each region are plotted by color. Interactive files showing the regions in the visualization are available [here](https://liangyy.github.io/brainxcan-docs/docs/example.html#63_Visualization_and_report).
